# Distinct Plasma Protein Profiles Distinguish Faster from Slower Disease Progression in HIV-1 and HIV-2 infections

**DOI:** 10.1101/2024.07.23.24310457

**Authors:** Emil Johansson, Jamirah Nazziwa, Eva Freyhult, Mun-Gwan Hong, Malin Neptin, Sara Karlson, Melinda Rezeli, Zacarias J. da Silva, Antonio J. Biague, Jacob Lindman, Angelica Palm, Patrik Medstrand, Fredrik Månsson, Hans Norrgren, Marianne Jansson, Joakim Esbjörnsson, the SWEGUB CORE group

**Affiliations:** Department of Translational Medicine, Lund University, Sweden; National Bioinformatics Infrastructure Sweden, Science for Life Laboratory, Department of Cell and Molecular Biology, Uppsala University, Uppsala, Sweden; Science for Life Laboratory, School of Engineering Sciences in Chemistry, Biotechnology and Health, KTH Royal Institute of Technology, Solna, Sweden; Department of Laboratory Medicine, Lund University, Lund, Sweden; BioMS − Swedish National Infrastructure for Biological Mass Spectrometry, Lund University, Lund, Sweden; National Public Health Laboratory, Bissau, Guinea-Bissau; Department of Clinical Sciences Lund, Lund University, Lund, Sweden; Nuffield Department of Clinical Medicine, University of Oxford, UK

**Keywords:** HIV-1, HIV-2, disease progression, plasma proteomics, tissue leakages, CD4+ T-cell level

## Abstract

The asymptomatic disease stage in HIV-2 infection is approximately twice as long compared to in HIV-1 infection, still the majority of HIV-2 infected individuals progress to AIDS in the absence of antiretroviral treatment. In this study, we applied data-independent acquisition mass spectrometry analysis of blood plasma samples collected from HIV negative, and HIV-1 or HIV-2 infected individuals in Guinea-Bissau with an estimated date of HIV infection, to explore associations between plasma proteome alterations and HIV disease progression. In total, 609 proteins were quantified and mapped towards publicly available data on tissue-enhanced genes, to provide insight on the tissue-specific origin of the detected proteins. Here we identified ten proteins that could differentiate between faster and slower HIV disease progression. The analysis also suggested a larger leakage of proteins from the sigmoid colon in HIV-1 compared to HIV-2 infection. Moreover, the levels of sigmoid colon and spleen tissue proteins were associated with disease progression among all HIV infected individuals. In conclusion, these results encourage further research on the role of both target and bystander cells in HIV disease progression.

## INTRODUCTION

Two types of related human immunodeficiency viruses (HIV) have been identified, HIV-1 and HIV-2. Previously, it was suggested that HIV-2 infection in most cases would not affect a normal lifespan, and that the majority of HIV-2 infected individuals did not progress to immunodeficiency and AIDS during follow-up (1-6). In contrast, we recently showed that the vast majority of HIV-2 infected most likely will progress to AIDS in the absence of antiretroviral treatment (ART) (7, 8). The median time from HIV infection to AIDS and HIV-related death were approximately twice as long among HIV-2 compared to HIV-1 infected individuals (7). However, and although these differences have been suggested to be partly attributed to a broader and more potent antiviral immune response among HIV-2 compared to HIV-1 infected individuals, the underlying molecular mechanisms behind the difference are unknown (9-16). In line with the slower disease progression, HIV-2 infected individuals typically have lower proviral and plasma viral load compared to HIV-1 infected individuals (1, 2, 17). Moreover, a large inter-individual difference in disease progression rate have also been observed between HIV-2 infected individuals – ranging from only a few years to more than two decades (7). The underlying mechanisms for this variation are also unknown. Importantly, an in-depth understanding of those disease-controlling mechanisms have the potential to unravel new targets relevant for therapeutics and vaccines against HIV (18).

Proteins represent the end-stage of gene expression, perform most of the work in cells, and are essential for the structure, function, and regulation of tissues and organs. Through recent technological advances in mass-spectrometry based proteomics, it is now possible to simultaneously identify and quantify hundreds of proteins from a sample volume of less than 10 µl (19). Even more intriguingly, through large-scale collaborative and open source initiatives like the Human Protein Atlas (HPA), it is now possible to create a bird’s eye-perspective of how proteome dynamics change in response to disease through in-depth comparisons of cell and tissue-specific proteomes between healthy and diseased conditions (20-23). Importantly, such comparisons may also unravel previously unknown key disease regulating proteins.

The overarching objectives of this study were i) to quantify the large-scale protein dynamics in blood plasma and associate this with HIV disease progression in general, and ii) to provide further insights on the pathogenic differences between HIV-1 and HIV-2 infection. The results show that the plasma levels of proteins released from both target and bystander cells can distinguish faster from slower HIV progressors.

## MATERIALS AND METHODS

### Study participants

Blood plasma samples were collected from HIV seronegative, and HIV-1 or HIV-2 infected study participants of a well-defined occupational cohort of police officers in Guinea-Bissau (7). In total, 225 individuals contracted HIV-1 and 87 individuals contracted HIV-2 following inclusion into the cohort (7). Of these individuals, 51 HIV-1 infected individuals and 44 HIV-2 infected individuals remained ART naïve and donated a plasma sample within the first three years following estimated date of infection (EDI). Based on sample availability, we included plasma samples collected from 25 HIV-1 infected, 25 HIV-2 infected, and 25 age-matched HIV seronegative individuals (see Table 1 for study participant characteristics).

**Table 1.**
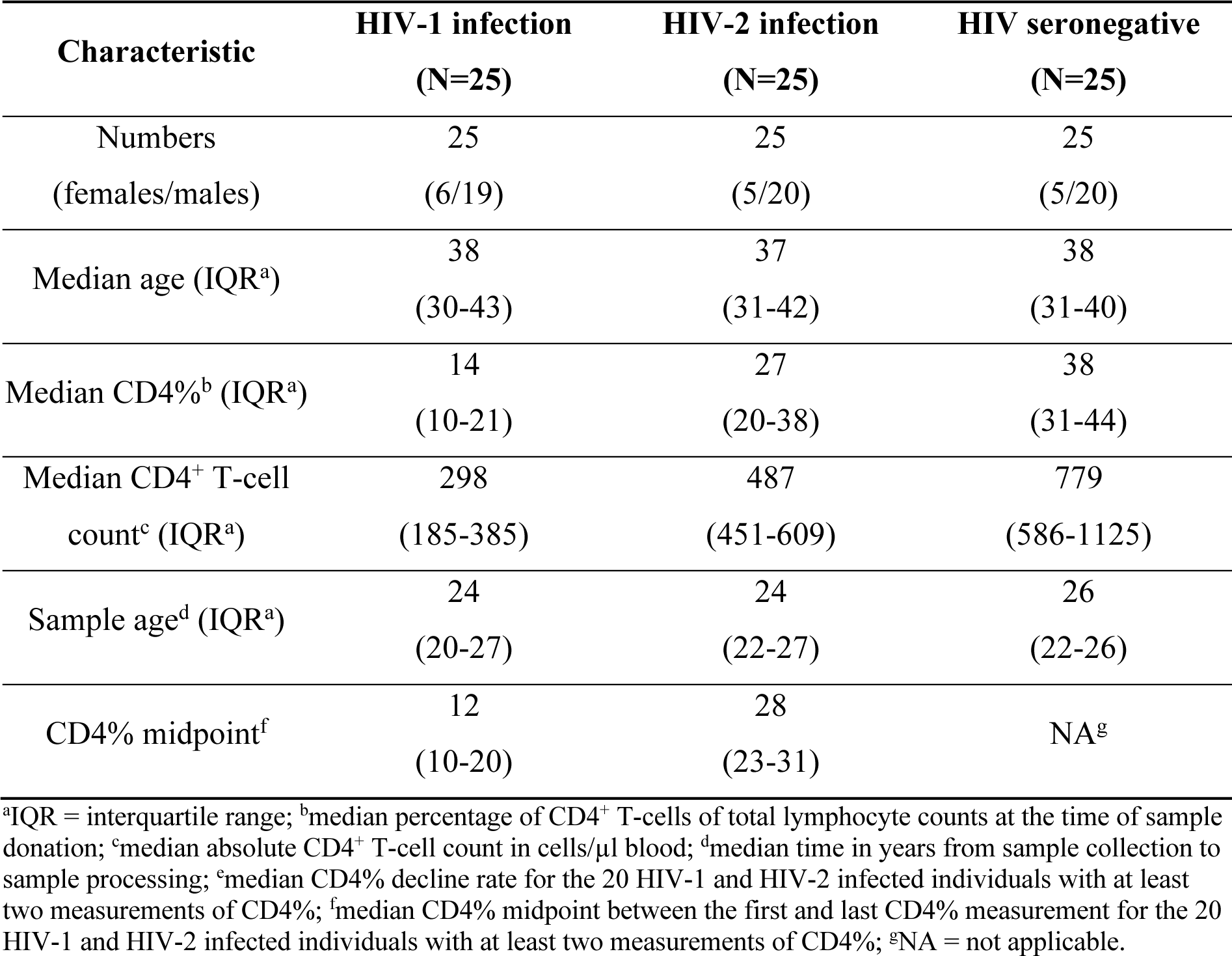
Characteristics of study participants.

### Ethics

The study was approved by the Ethical committee at Lund University, and the National Ethical Committee, Ministry of Public Health in Guinea-Bissau. Informed consent was obtained from all participants. All samples used in this study were collected according to the guidelines in the Declaration of Helsinki.

### Sample collection and processing

Plasma was collected, and the absolute CD4+ T cell count and percentage of CD4+ T cells of lymphocytes (CD4%) were measured as previously described (7, 24, 25). Prior to the sample preparation for the nano-scale liquid chromatography tandem mass spectrometry (nLC-MS/MS) analysis, the 14 most abundant proteins (albumin, alpha-1-acid glycoprotein, alpha-1-antitrypsin, alpha-2-macroglobulin, apolipoprotein A1, fibrinogen, haptoglobin, and transferrin, and the kappa and lambda light chains of immunoglobulin (Ig) G, IgA, IgM, IgD, and IgE) were depleted from four microlitre plasma using the High Select™ Top14 Abundant Protein Depletion Mini Spin Columns (ThermoFisher Scientific) according to the manufacturer’s instructions. The depleted samples were thereafter prepared using a filter-aided sample preparation approach. Briefly, the depleted plasma was transferred to Amicon 10 kDa filters (Merck Millipore), concentrated, denatured in 8 M urea (Merck), reduced in 35 mM dithiothreitol (Merck) for one hour at 37 °C, and alkylated with 55 mM iodoacetamide (Merck) for 30 minutes at room temperature in the dark.

The proteins were subsequently washed twice with 100 mM ammonium bicarbonate before elution in 100 mM ammonium bicarbonate. The protein concentration was determined using Qubit 4 Fluorometer (ThermoFisher Scientific) and the Invitrogen Qubit Protein BR Assay Kit (ThermoFisher Scientific). A final concentration of one µg/µl of sequencing Grade Modified Trypsin (Promega) was added to 30 µg protein, and the samples were digested at 37 °C for 16 hours. The digestion was stopped using 10% trifluoroacetic acid and the peptide concentration was thereafter determined using the Nanodrop 2000 spectrophotometer (ThermoFisher Scientific). Finally, the sample were store at -80 °C until nLC-MS/MS analysis.

### Mass spectrometry analysis and data extraction

Prior to the nanoflow liquid chromatography (nLC)-MS/MS analysis, samples were spiked with indexed retention time (iRT) peptides. One µg of peptide solution per sample were injected onto the nLC column. A Dionex Ultimate 3000 Rapid Separation LC (RSLC) nano Ultra-performance LC system coupled to an Orbitrap Exploris 480 MS with FAIMS Pro interface (Thermo Scientific) was used for the nLC-MS/MS analysis. The peptides were loaded onto an Acclaim PepMap 100 C18 (75 μm × 2 cm, 3 μm, 100 A, nanoViper) trap column and separated on an Easy-spray PepMap RSLC C18 column (75 μm × 50 cm, 2 μm, 100 A) (Thermo Scientific) using a flow rate of 300 nL/min, and a column temperature of 60 °C. A 90 min gradient was applied for separation, using solvents A (0.1% formic acid) and B (0.1% formic acid in 80% Acetonitrile (ACN)), increasing solvent B from 5% to 25% in 75 min, then to 32% in the next 9 min, and to 45% in following 6 min. Finally, the gradient increased to 95% solvent B for 2 min, and then continuing at 95% for another 5 min.

For DIA, full MS resolution was set to 120 000 (at 200 m/z), AGC target value was 300% with a maximum injection time (IT) of 45 ms. Full MS mass range was set to m/z 380-1100. MS2 scans were acquired with a resolution of 30 000 (at 200 m/z), fragmentation with a normalized collision energy (NCE) of 32, AGC target value was set to 1000% with automatic IT. Twenty-six variable isolation windows of 35.0, 22.5, 35.0 and 50.0 Da were used with an overlap of 0.5 Da, and with 1, 20, 1 and, 4 loop counts, respectively. For FAIMS, two compensation voltages (CVs) of -45 and -60 V were used with a carrier gas flow of 3.5 L/min and the spray voltage was set to 2.1 kV.

For data dependent acquisition (DDA) experiments (library generation), full MS resolution was set to 120 000 (at 200 m/z), AGC target value was 300% with a maximum IT of 45 ms. Full MS mass range was set to 350-1400 m/z. MS2 scans were acquired with a resolution of 30 000 (at 200 m/z), fragmentation with a NCE of 30, AGC target value was set to 100% with automatic IT. The precursor isolation window was set to 1.6 m/z, intensity threshold was kept at 2e4, and 45 s dynamic exclusion was used. For FAIMS CVs of -45 and -60 V were used with cycle times of 1.7 and 1.3 s, respectively.

### Construction of spectral library for the DIA-MS Analysis

We created a combined spectral library from DIA measurements of the individual plasma samples, and from DDA measurements of a sample pooled from 19 depleted plasma samples and fractionated by off-line high-pH reversed-phase chromatography (Pierce High pH Reversed-Phase Peptide Fractionation Kit, Thermo Fisher Scientific). The spectral library was generated using the Pulsar search engine incorporated into Spectronaut 16.0.220606.53000 (Biognosys, Schlieren, Switzerland) according to the factory default settings. All generated raw data were searched against the UniProtKB reviewed human database, including isoforms (accessed on May 27, 2022; 42,363 entries) combined with the HIV-1 and HIV-2 reference proteomes (accessed on May 27, 2022; 189 entries) (37). The parameters included trypsin as digestion enzyme, two missed cleavages allowed, carbamidomethyl cysteine as fix modification, and methionine oxidation and acetylation on protein N-terminal as variable modifications. The number of identifications was controlled by false discovery rate (FDR) of 1% at peptide, and protein level, respectively. The spectral library consisted of 23 581 precursors, 16 075 peptides, 2 427 proteins, and 1 510 protein groups.

### DIA-MS targeted data extraction

DIA data files were searched against the spectral library using the Biognosys (BGS) factory default settings in Spectronaut 16.0.220606.5300 (Biognosys, Schlieren, Switzerland). The number of identifications were filtered at an FDR of 1% at both peptide and protein levels.

### Data processing and quality control

Protein levels were determined from peptide precursor and fragment ion intensities using the *iq* package (v1.9.6) in R (v4.2.1) (26, 27). The quantification data for the remains of the 14 top abundant proteins were excluded from the data set, as they could not be reliably quantified after the depletion. Proteins with more than 20% missingness were excluded from the analysis, as is custom in the field (28). For the remaining proteins with missing values, a random intensity value between one and the lowest quantified value was imputed to the samples with missing quantification. Eight normalization methods were evaluated using the NormalyzerDE package (v1.16.0), and variance stabilizing normalization (VSN) was found to perform best for this dataset. The protein levels were thereafter normalised by the VSN method using the VSN package (v3.66.0).

### Statistical analyses

#### Disease progression rate analysis

Disease progression rate in HIV-2 infected individuals was evaluated by the CD4% at time of sample donation and by the CD4% midpoint, as previously described (29, 30). Briefly, the CD4% midpoint value was defined as the midpoint of a regression line fitted to the CD4% measurements obtained from each individual. These two markers have previously been found to best distinguish faster from slower HIV-2 disease progressors (29).

#### Data quality control

The NormalyzerDE package (v1.16.0) was used with default settings to determine which normalisation algorithm to be used (31). The dimensionality of the protein abundance data reduced by performing a principal component analysis (PCA) using the *FactoMineR* package (v2.6) in R (32, 33). The association between age, sex and HIV status of the study participant, CD4% and absolute CD4+ T cell count at sample date, and sample age with the first five components was evaluated by a Pearson correlation test using the *stats* package (v4.2.1) (33) in R.

#### Differential protein level analysis

Normalised protein intensity values were compared between the three different HIV status groups by performing linear regression analyses with sample age as a covariate in the regression model, using the *limma* packages (v3.54.0) in R (34). The obtained p-values were corrected for multiple testing according to the Benjamini-Hochberg’s (BH) method using the *limma* package in R. Adjusted p-values <0.05 were considered significant.

#### KEGG pathway analysis

Kyoto Encyclopedia of Genes and Genomes (KEGG) pathway enrichment analysis was performed using the *clusterProfiler* package (v4.6.0) in R (35, 36). The p-values were adjusted according to the BH method and adjusted p-values <0.05 were considered significant.

#### Tissue damage analysis

To assess protein expression associated with specific tissues, a previously described tissue-enriched transcriptional signature dataset created using the Genotype-Tissue Expression (GTEx) database was used (37). In short, converted GTEx-acquired read counts were converted to trimmed mean of M values, the values were normalized to z-scores for each gene across tissues, and genes with a z-score above three were considered tissue enriched. To calculate the tissue scores, the protein intensity was first adjusted for sample age. The adjusted plasma protein dataset was then subset to only include the proteins identified as tissue enriched. The intensity values were converted into z-scores based on the mean protein value across all donors. The individual tissue scores for each donor were defined as the median z-score value of the proteins in each tissue gene list.

#### Kaplan-Meier analysis

Kaplan-Meier analysis was used to assess time to AIDS, as previously described (7), using the *survminer* (0.4.9) and *survival* (3.5-0) packages in R (38, 39). In brief, AIDS was defined according to WHO staging as CD4 counts of ≤200 cells/μL, or CD4% of ≤14%; or AIDS-defining symptoms. Cases that did not reach AIDS during follow-up were right censored at their last visit. Statistical comparisons were done using the log-rank test.

#### Data visualisation

The *ggplot2* package (v3.4.0) was used to produce the plots displaying PCA coordinates, violin plots displaying disease progression values and tissue scores in R (40). The volcano plots were produced using the *EnhancedVolcano* package (1.16.0) and the ridge plots displaying enrichment of KEGG pathways using the *enrichplot* package (v1.18.3) in R (35, 36, 41).

#### Code and data availability

Upon publication all codes will be made available at the systems virology GitHub website and the data will be uploaded to an appropriate repository.

## RESULTS

### Study participants

To investigate differences between chronic HIV-1 and HIV-2 infections, archived plasma samples donated within three years from the estimated date of infection from ART naïve HIV-1 (n=25) and HIV-2 (n=25) infected individuals were analysed by DIA-MS. In addition, plasma samples from HIV seronegative (n=25) individuals were included as controls. For 20 HIV-1 (80%) and 20 HIV-2 infected (80%) individuals, additional CD4% measurements had been collected, which allowed us to compare the disease progression rate of these individuals with the rest of the cohort. No selection regarding disease progression rate or age was observed for the selected study participants as compared to the whole cohort (Figure S1). Further characteristics of the study participants are described in Table 1.

### HIV-1 and HIV-2 infections induce increased release of proteins involved in pathogen responses

VSN normalisation algorithm was chosen by NormalyzerDE package as it showed the lowest values of intragroup coefficients of variation and median absolute deviation of all replicate groups by HIV status (Figure S2A). In total, 1 334 unique plasma proteins were detected. Following the exclusion of protein with more than 20% missingness across all samples, as well as proteins targeted by the depletion kit, 609 protein (46%) were included for subsequent analyses. The quality control indicated that age, sex, sample preparation order, or order of nLC-MS/MS analysis (running order) were not associated with any clear distortion or skewing of the raw data intensity (Figure S2B). However, sample age (the time between sample collection and experiment) was found to be associated with PC1. In addition, the intensities of 161 proteins were found to be associated with sample age in a linear regression model. We therefore included sample age as a covariate in our linear regression model, and adjusted protein intensity for sample age before generating z-scores for further analysis of the impact of infection on cells and tissues. Compared to the HIV seronegative individuals did HIV-1 infected individuals have increased levels of 67 proteins and decreased levels of 53 proteins, and HIV-2 infected individuals had increased levels of 32 proteins and decreased levels of 21 proteins (Figure 1A). Forty-five of the 173 differentially expressed proteins were found among both HIV-1 and HIV-2 infected individuals. Both HIV-1 infected and HIV-2 infected individuals displayed enrichment of the human papilloma virus infection, ECM-receptor interaction, focal adhesion, and phagosome KEGG pathways (Figure 1B). In addition to the shared KEGG pathways, an enrichment of PI3K-Akt signalling and a decrease of the glycolysis/gluconeogenesis pathway were observed in HIV-1 infection, whereas an enhancement of the neutrophil extracellular trap formation and Malaria pathways were seen in HIV-2 infection (Figure 1B). Taken together, these findings indicate that chronically HIV-1 and HIV-2 infected individuals display overall similar increase in proinflammatory processes, but with some virus type-specific alterations.

**Figure 1.**
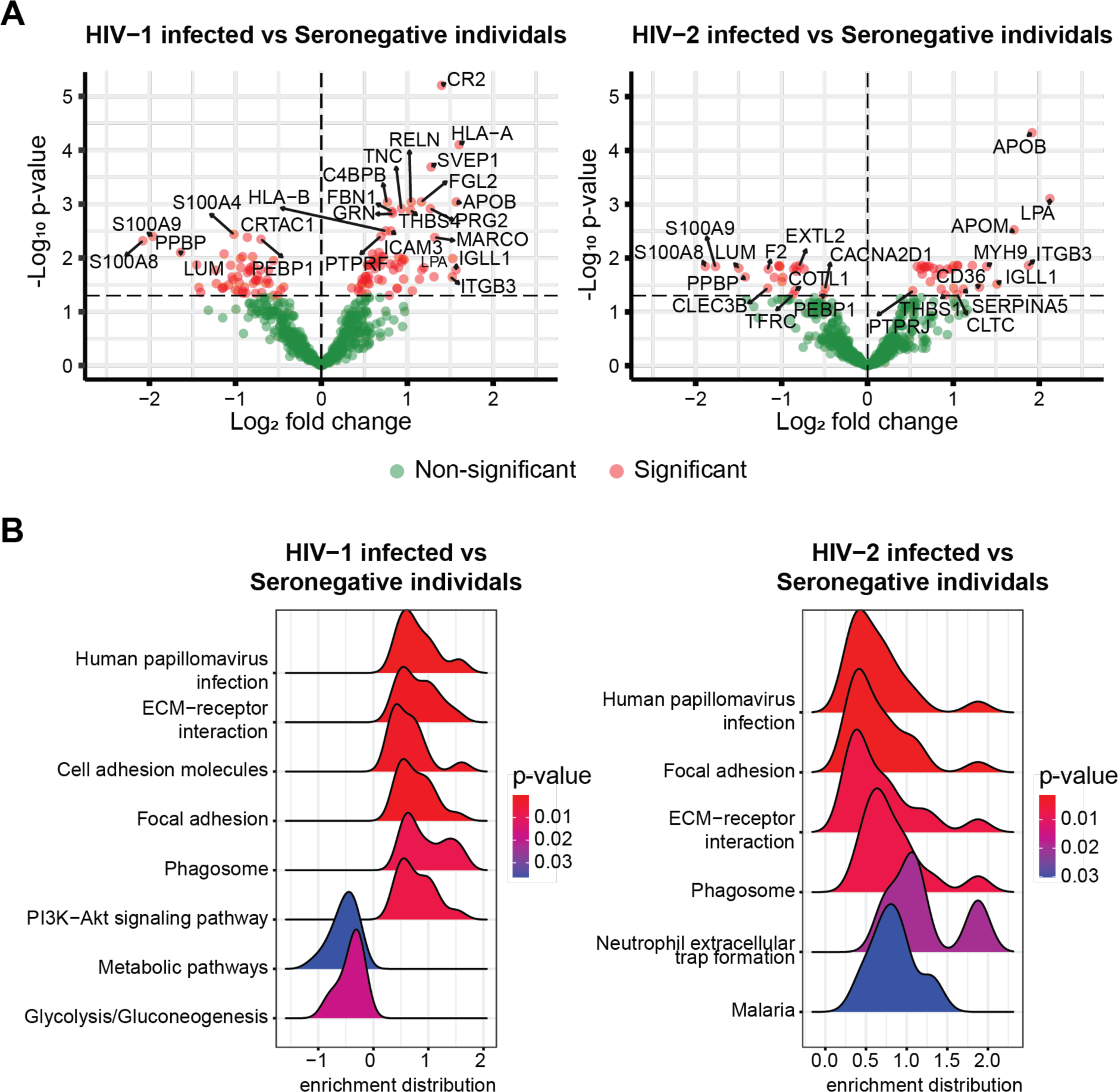
HIV-1 and HIV-2 induce similar alterations of the blood plasma proteome. A) A linear regression model was used to identify differentially expressed proteins among HIV-1 and HIV-2 infected individuals compared to seronegative individuals (Benjamini-Hochberg [BH] adjusted, p<0.05). B) KEGG pathway analysis was used to identify significantly up or downregulated biological processes of the detected plasma proteins (BH adjusted p<0.05).

### Plasma levels of both target and bystander cell-derived proteins are associated with disease progression

To delineate the impact of HIV-1 and HIV-2 infection on different cell types, we utilised the HPA resource to define proteins with cell type enriched expression patterns. The analysis indicated that 348 of the 609 detected proteins were cell type enriched. Of these, the levels of 86 proteins differed significantly between any of the three HIV status groups (Figure S3). To identify cell type enriched proteins associated with disease progression, we next investigated the correlation between protein intensity and CD4% and CD4% midpoint. We found that ADA2 (enriched among macrophages), TNC (smooth muscle cells), NRP2 (endothelial cells), RAD23B (late spermatids), and LGALSL (photoreceptor cells and keratinocytes) were significantly associated with both CD4% levels (r=-0.571; p<0.001, r=-0.516; p<0.001, r=-0.510; p<0.001, r=0.543; p=0.001, and r=0.363; p=0.003, respectively), and CD4% midpoint (r=-0.499; p<0.001, r=-0.463; p<0.001, r=-0.413; p=0.002, r=0.561; p=0.004, and r=0.354; p=0.022, respectively) when all HIV infected individuals were analysed together (Figure 2). In addition, ADA2, TNC, NRP2, and RAD23B were associated with CD4% levels when HIV-2 infected individuals were analysed separately (r=-0.603; p<0.001, r=-0.522; p=0.002, r=-0.483; p=0.006, and r=0.641; p=0.012, respectively). Taken together, these results indicated that HIV-1 and HIV-2 disease progression was associated with dysregulation of wide variety of cell types.

**Figure 2.**
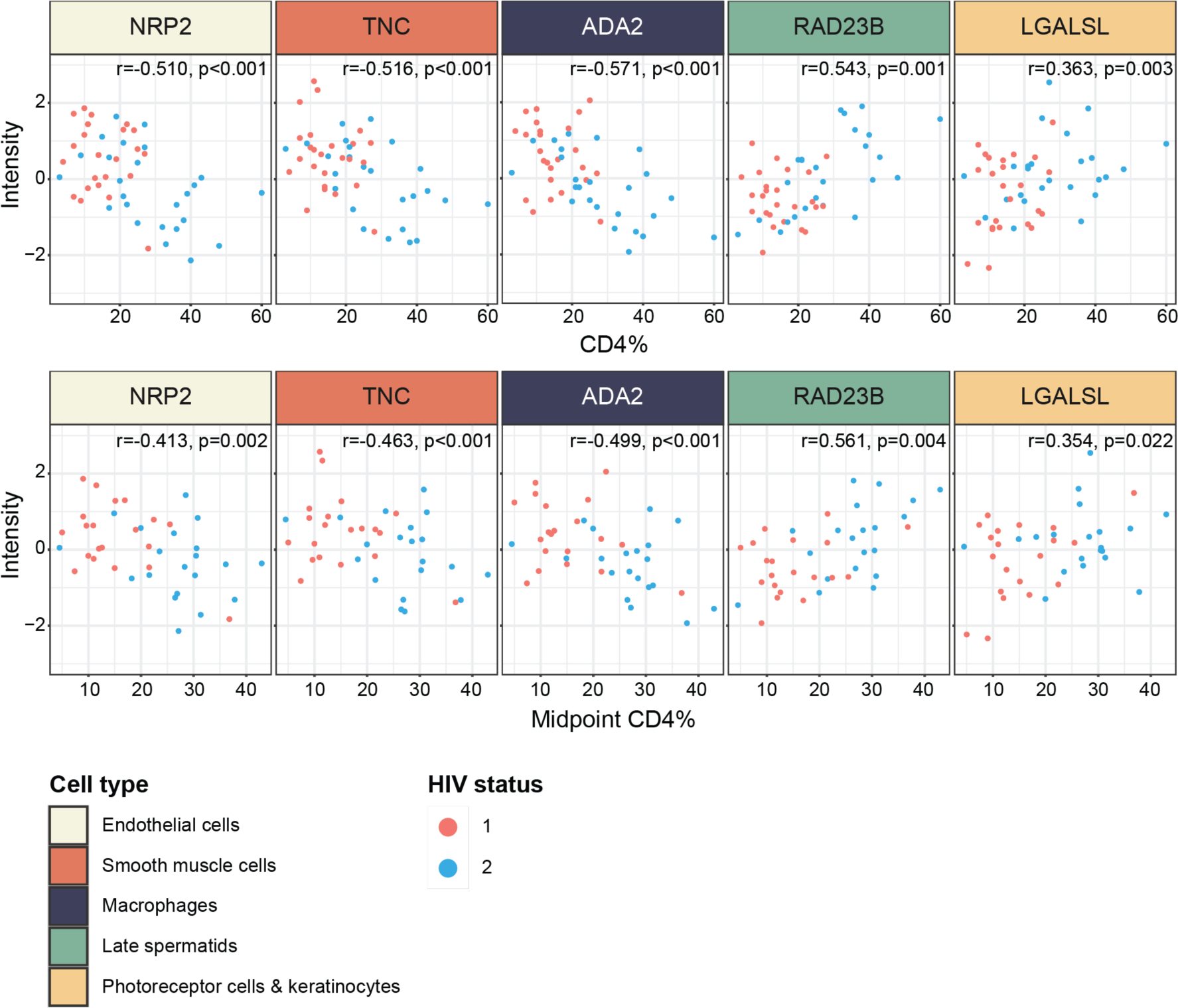
Proteins released from macrophages and non-haematopoietic cells correlate with disease progression. Pearson correlation test followed by BH correction for multiple testing identified five proteins significantly (BH adjusted p<0.05) associated with CD4% (upper row) and CD4% midpoint (lower row). Protein intensities reflect z-score normalised protein intensities.

### HIV infection induce colon and spleen tissue engagement

To further disentangle the tissue-origin of the proteins in plasma that were associated with disease progression, we used a previously described tissue-specific transcriptional signature dataset created using the GTEx database to generate protein tissue scores (37). In total, 359 of the 609 detected proteins overlapped with the tissue-specific transcriptional signature dataset. The HPA resource provides information about the predicted location of proteins, including proteins predicted to be actively secreted or released to blood (hereafter referred to as secreted plasma proteins) (42). The remaining proteins detected in plasma are expected to have leaked into the blood from tissues or dying cells (hereafter referred to as leakage proteins). We therefore divided the dataset into blood cell-derived and leakage proteins to gain a deeper insight into associations between plasma proteins and HIV infection. To increase the likelihood of identifying signs of tissue damage related to activation or death of bystander cells, we removed proteins enriched in whole blood cells from the tissue-specific transcriptional signature dataset (37). This resulted in 36 tissue scores from proteins not secreted to blood, and 28 tissue scores from proteins secreted to blood (Figure S4). The total number of proteins in each tissue ranged from 1-18 for proteins not secreted to blood, and 1-89 for proteins previously designated as secreted to blood. Among the leakage protein-derived tissue scores, HIV-1 infected individuals had significantly higher sigmoid colon tissue score than HIV-2 infected (p=0.002) and HIV seronegative individuals (p<0.001) (Figure 3A, Table 2). The sigmoid colon tissue score also correlated with both CD4% levels (r=-0.560, p<0.001), and midpoint CD4% (r=-0.480, p=0.009) among all HIV infected individuals (Figure 3B). In addition, adrenal gland, oesophagus mucosa, oesophagus muscularis, nerve tibial, and liver tissue scores differed significantly between at least two of the HIV status groups (Figure S5A). However, the tissue scores did not correlate with CD4% levels nor midpoint CD4%.Among the plasma protein-derived tissue scores, HIV-1 and HIV-2 infected individuals had significantly higher spleen tissue scores compared to HIV seronegative individuals (p=0.004 and p=0.044, respectively, Figure 3C, Table 2). The spleen tissue score also correlated with CD4% (r=-0.386, p=0.045) among all HIV infected individuals (Figure 3D). In addition, spinal cord, transverse colon, oesophagus mucosa, heart left ventricle, pancreas, testis, and thyroid tissue scores differed significantly between at least two of the HIV status groups, but were not associated with CD4% (Figure S5B). Altogether, these results suggest that increased release of proteins from the sigmoid colon and spleen was associated with disease progression, assessed by lower CD4%.

**Figure 3.**
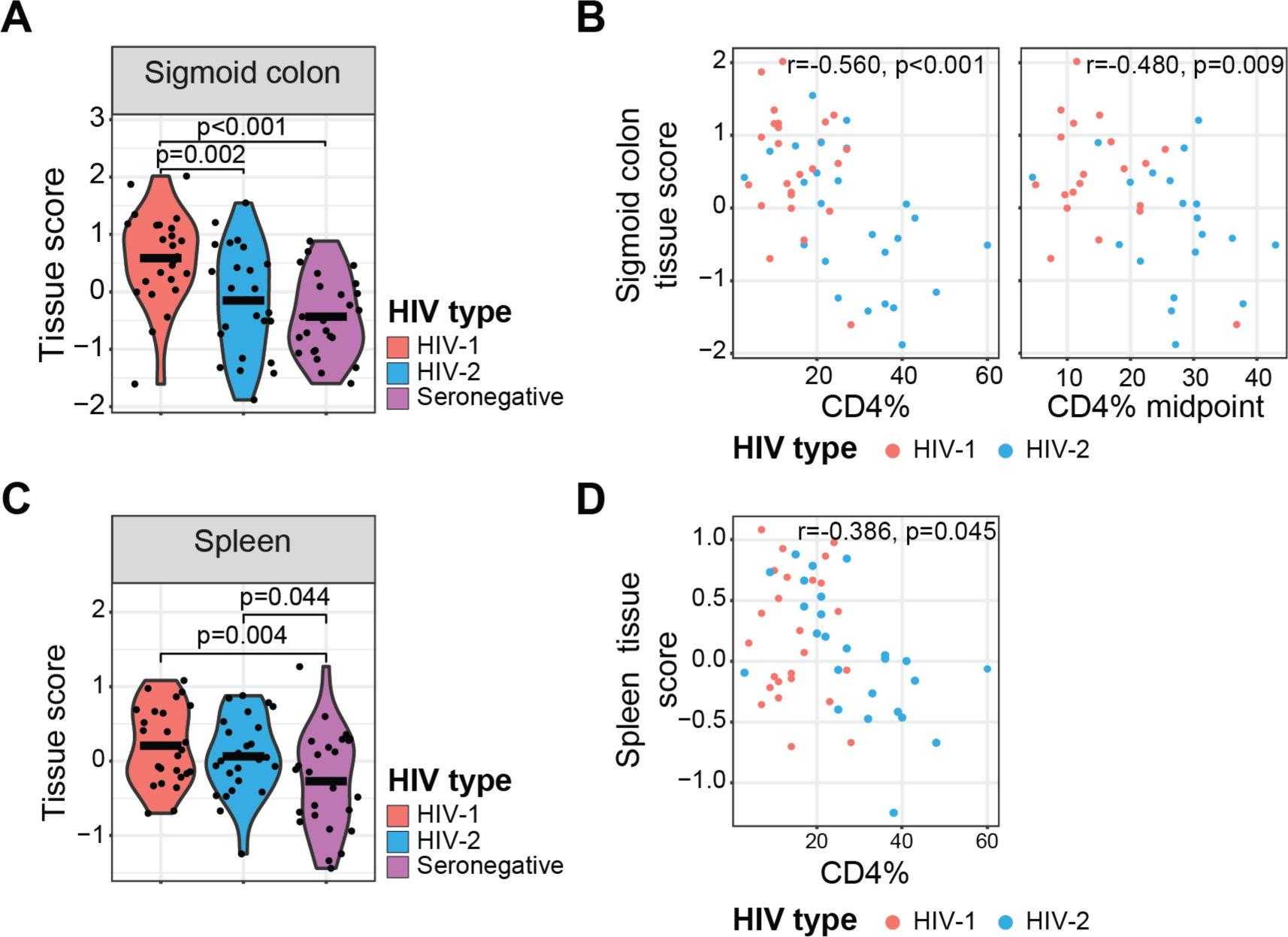
HIV-1 induces plasma elevation of proteins derived from tissue associated with disease progression. A) Group-wise comparison of sigmoid colon tissue score, consisting of leakage proteins, and B) correlation between sigmoid colon tissue scores and CD4% and CD4% midpoint. C) Group-wise comparison of spleen tissue score, consisting of secreted proteins, and D) correlation between spleen tissue score and CD4%. Tissue scores represent the median z-score normalised protein level of all proteins enriched in sigmoid colon or spleen. Statistically significant group-wise differences were determined by ANOVA, followed by Benjamini-Hochberg (BH) correction for multiple testing (p<0.05). Statistically significant correlations were determined using the Pearson correlation test followed by BH correction for multiple testing (p<0.05). Mean values are depicted in violin plots.

**Table 2.** Tissue scores stratified by HIV status groups.

| **Tissue** | **HIV-1 infection**<br>**(N=25)** | **HIV-2 infection**<br>**(N=25)** | **HIV seronegative**<br>**(N=25)** |
| --- | --- | --- | --- |
| Sigmoid colon<sup>a</sup> | 0.585<br>(0.802) | -0.152<br>(0.926) | -0.433<br>(0.694) |
| Spleen<sup>a</sup> | 0.208<br>(0.525) | 0.063<br>(0.525) | -0.271<br>(0.666) |

<sup>a</sup>Mean tissue score (standard deviation).

### Plasma levels of bystander cell-derived protein are associated with CD4%

To delineate which cells that drive the observed difference in tissue scores described above, we compared the levels of the proteins included in each of the tissue scores found to differ between at least two of the HIV status groups. We found that the levels of the leakage proteins NRP2, TNC, and GGH were higher among HIV-1 compared to both HIV-2 infected (p=0.002, p=0.012, and p<0.001, respectively), and HIV seronegative individuals (p=0.001, p<0.001, and p=0.001, respectively, Figure 4A, Table 3). In addition, HIV-1 infected individuals had higher GOLM1 (p=0.002), and lower SH3BGRL2 and GSTO1 levels (p=0.009 and p=0.002, respectively), whereas HIV-2 infected individuals had higher ACTN1 levels (p=0.012), compared to HIV seronegative individuals. Moreover, a negative correlation between NRP2 levels and both midpoint CD4% (r=-0.463, p<0.001) and CD4% (r=-0.510, p<0.001) was observed analysing HIV infected individuals together (Figure 4B), whereas TNC (r=-0.516, p=0.001), SH3BGRL2 (r=0.410, p=0.017), GGH (r=-0.386, p=0.022), GSTO1 (r=0.348, p=0.036), GOLM1 (r=-0.342, p=0.036), and ACTN1 (r=0.340, p=0.036) levels were associated with CD4% levels when analysing HIV infected individuals together (Figure 4B). Nineteen additional proteins, nine leakage, and ten secreted proteins, differed significantly between at least two HIV status groups, but were not associated with midpoint CD4%, or CD4% level (Figure S6).

**Figure 4.**
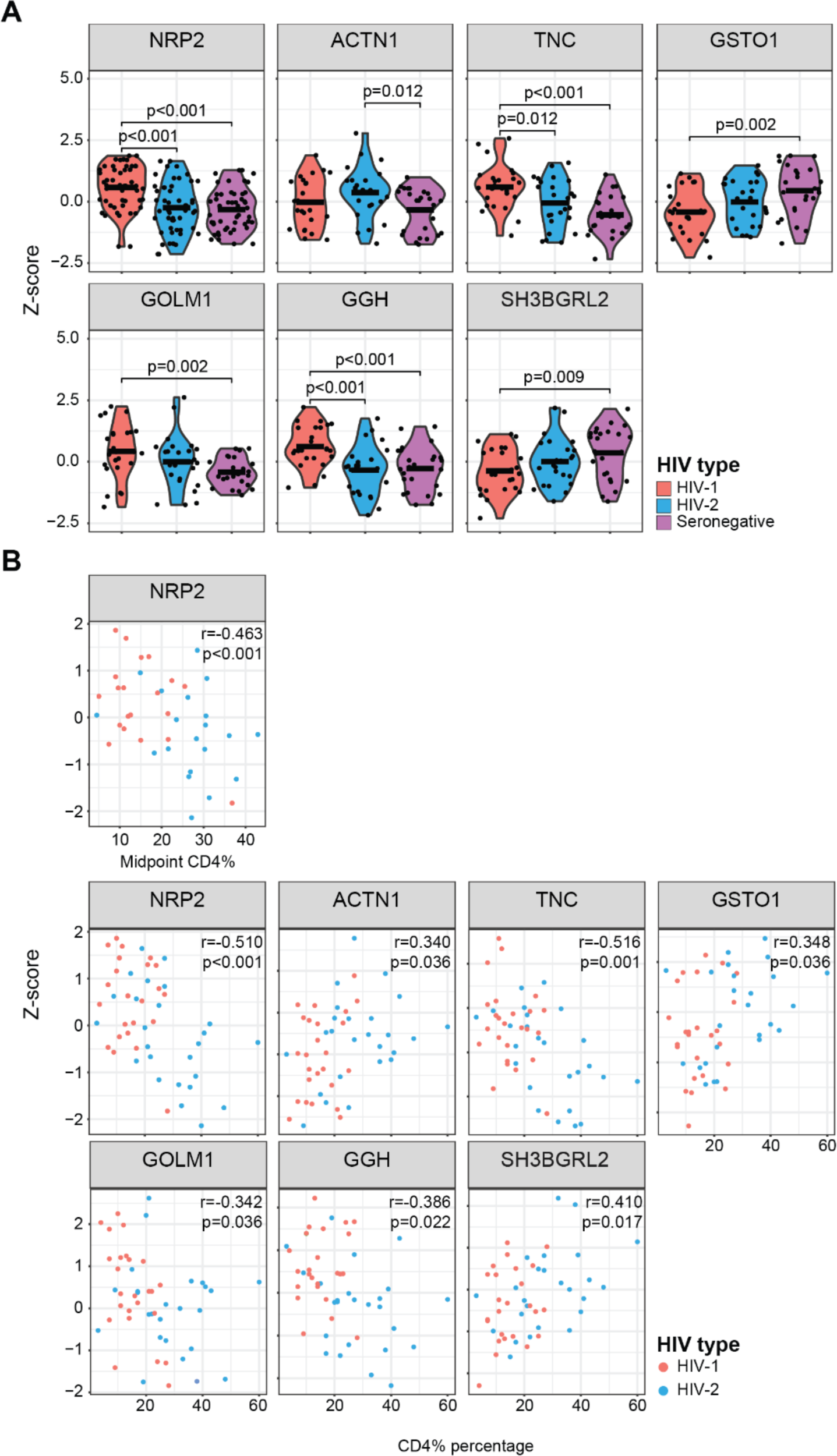
Plasma levels of seven differentially expressed proteins are significantly associated with CD4%. A) Group-wise comparison of the z-score normalised protein intensity and B) correlation to CD4% and CD4% midpoint. Statistically significant group-wise differences (BH adjusted p<0.05) were determined by ANOVA followed by BH correction for multiple testing. Statistically significant correlations (BH adjusted p<0.05) were determined using the Pearson correlation test followed by BH correction for multiple testing. Mean values are depicted in violin plots.

**Table 3.** Levels of proteins associated with disease progression and stratified by HIV status.

| **Tissue** | **HIV-1 infection**<br>**(N=25)** | **HIV-2 infection**<br>**(N=25)** | **HIV seronegative**<br>**(N=25)** |
| --- | --- | --- | --- |
| NRP2<sup>a</sup> | 0.573<br>(0.888) | -0.250<br>(1.020) | -0.323<br>(0.832) |
| ACTN1<sup>a</sup> | -0.026<br>(0.992) | 0.368<br>(1.030) | -0.342<br>(0.884) |
| TNC<sup>a</sup> | 0.597<br>(0.907) | -0.054<br>(0.980) | -0.543<br>(0.786) |
| GSTO1<sup>a</sup> | -0.427<br>(0.914) | -0.013<br>(0.902) | 0.441<br>(1.020) |
| GOLM1<sup>a</sup> | 0.427<br>(1.110) | -0.002<br>(1.070) | -0.425<br>(0.570) |
| GGH<sup>a</sup> | 0.616<br>(0.843) | -0.340<br>(0.993) | -0.277<br>(0.890) |
| SH3BGRL2<sup>a</sup> | -0.374<br>(0.888) | 0.010<br>(0.953) | 0.364<br>(1.050) |

<sup>a</sup>Mean tissue score (standard deviation).

Next, we used publicly available single-cell RNA sequencing (scRNA seq) data sets available from the HPA resource to determine the cell types associated with the proteins that were linked to disease progression. Although NRP2 was included in both the sigmoid colon and oesophagus muscularis tissue-specific transcriptional signatures, the expression was only detected in oesophagus tissue of the available scRNA seq data sets. Within the oesophagus tissue scRNA seq data set, NRP2 and SH3BGRL2 expression was found to be enriched among endothelial cells, while ACTN1 expression was enriched among both endothelial and smooth muscle cells (Figure S7A). The expression of GOLM1 was included in the tissue-specific transcriptional signatures. However, no adrenal gland scRNA seq dataset is available through the HPA resource, which prevented us from identifying which cell types that express GOLM1 in the adrenal gland. Instead, GOLM1 expression was highest in a cluster of mixed cell types in pancreatic tissue (Figure S7B). Although TNC expression was included in the tissue-specific transcriptional signature, it was not detected in colon, small intestine, or rectum scRNA seq datasets available through the HPA resource. Instead, TNC expression was highest in smooth muscle cells within the thymus scRNA seq dataset (Figure S7C). Of note, no smooth muscle cells were detected in the intestine or colon scRNA seq dataset. Within the liver scRNA seq data set, GSTO1 and GGH expression was enriched among hepatocytes (Figure S7D). Taken together, these findings indicate that both HIV-1 and HIV-2 infection impact bystander cells, such as endothelial cells, smooth muscle cells, fibroblasts, and hepatocytes, to release proteins into plasma, and that these engagements are associated with HIV disease progression.

### Identification of plasma proteins that differentiate slower from faster HIV disease progressors

To further assess the impact of the ten proteins that correlated with CD4% midpoint and/or CD4% levels, we used unsupervised hierarchical clustering to determine groups associated with expression of these proteins. The analysis suggested two different clusters (Figure 5A), whereof the smaller cluster of 13 study participants (cluster 1) was characterised by lower ADA2, GGH, NRP2, and TNC levels compared to the study participants in the other cluster (cluster 2, N=37). Two study participants from cluster 1, and eight from cluster 2 had not been followed longitudinally. When comparing the time to AIDS onset in the remaining 40 individuals, we found a significant difference in time to AIDS between the two groups (p=0.006, Figure 5B). As only two individuals from cluster 1 progressed to AIDS during the 17.6 years follow-up period, the median time to AIDS could not be determined for this cluster. The median time to AIDS was 3.5 years (95% confidence interval 2.9-8.1 years) for individuals in cluster 2. In conclusion, we identified ten proteins, released from both target and bystander cells, that could be used to group HIV infected individuals into groups with different disease progression rates. These findings highlight the impact of not only target cell damage, but also the impact of HIV-1 and HIV-2 infections on bystander cell engagement on HIV disease progression.

**Figure 5.**
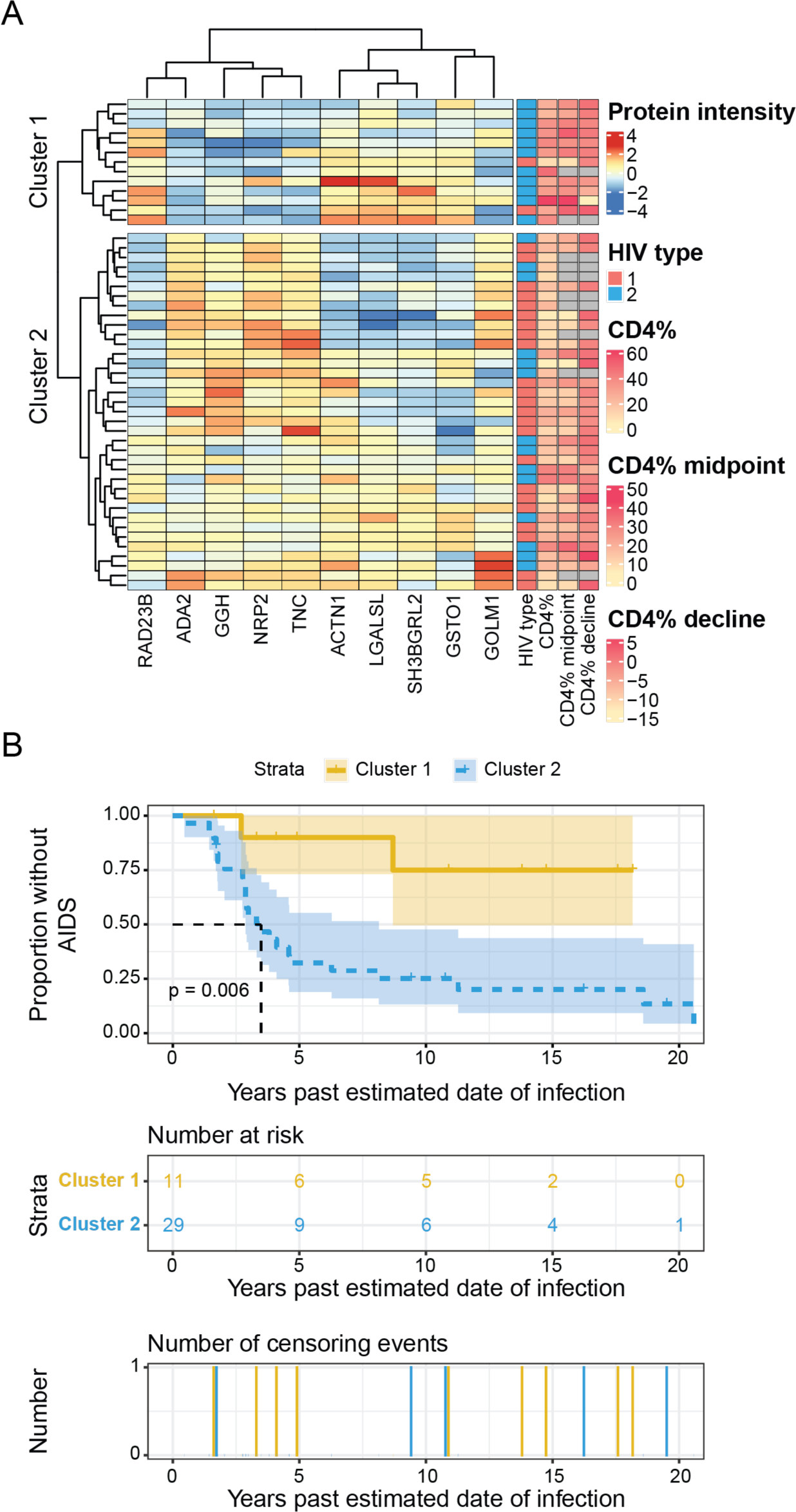
Faster and slower HIV progressors are separated based on plasma levels of ten proteins. A) Hierarchical clustering of study participants (row dendrograms) based on the expression of RAD23B, ADA2, GGH, NRP2, TNC, ACTN1, LGALSL, SH3BGRL2, GSTO1, and GOLM1. The heatmap displays z-score normalised protein intensity. HIV status, CD4% at time of sample collection (CD4%), midpoint CD4% between the first and last CD4% measurement (midpoint CD4%), and CD4% decline rate between the first and last CD4% measurement (CD4% decline) are illustrated to the right of the heatmap for each participant. Participants lacking follow-up data for CD4% are noted with a grey box for midpoint CD4% and CD4% decline. B) Kaplan-Meier curves of time to AIDS for individuals in the two defined clusters of study participants. Tick marks indicate time of censoring of participant. Log rank p-value is reported.

## DISCUSSION

In this study, we have identified ten proteins in plasma that could be used to distinguish faster from slower HIV progressors using an unsupervised hierarchical clustering approach. Interestingly, apart from ADA2, the remaining nine proteins have not been associated with active secretion into the blood. This suggests that increased cell death or leakage from tissues may distinguish faster from slower HIV progression. In addition, the ten proteins have all been described to be expressed in a range of different types of cells (43, 44). Although the main targets of both HIV-1 and HIV-2 are CD4+ T cells and myeloid cells (45), it is well-known that HIV-1 can cause perturbations in several cell types and organs, such as non-haematopoietic cell types like endothelial cells, fibroblasts and smooth muscle cells, both through direct and indirect modes of action (46, 47). Moreover, this dysregulation of bystander cells can impact the function of several organs, such as the gastrointestinal tract (GIT) where HIV-1 infection has been shown to cause both epithelial cell death and to increase epithelial permeability (48, 49). Disruption of the intestinal epithelium is accompanied by an increased microbial translocation, which has been suggested to play a key role in the exhaustion of the immune system during both HIV-1 and HIV-2 infection (48, 50, 51). Indeed, plasma levels of LPS have been reported to increase during disease progression in both infections (51). However, and in contrast to HIV-1 infection, HIV-2 and non-pathogenic SIV infections have been reported to have a less pronounced effect on the epithelial integrity (48, 52). In line with this, we observed statistically significant elevated blood levels of leakage proteins from the sigmoid colon among HIV-1, compared to HIV-2 infected and HIV seronegative individuals. In further agreement with the association between GIT damage and HIV-1 and HIV-2 disease progression, we found that the sigmoid colon tissue score was associated with the two disease progression markers CD4% at time of sampling and CD4% midpoint during follow up when analysing HIV-1 and HIV-2 infected individuals together.

The association between proteins leaked from sigmoid colon and secreted from the spleen, two known sites of HIV replication, indicates that replication in these compartments may play a key role in both HIV-1 and HIV-2 disease progression (53). Although the level of plasma viral load (pVL) in HIV-2 infected individuals has been suggested to be associated with disease progression rate (2), HIV-2 infected individuals has still been shown to progress in the absence of viraemia (24, 54). Further, the pVL of viraemic HIV-2 infected AIDS patients has been found to be lower than that of HIV-1 infected AIDS patients (55), and HIV-2 infected individuals have been found to develop AIDS at higher CD4% levels than HIV-1 infected individuals (7, 55, 56). This suggests that HIV-2 disease progression is not solely dependent on pVL, but it is still not known whether HIV-2 disease progression is driven by continuous HIV-2 replication and exposure to HIV-2 antigens, or due to persistent inflammation observed also in HIV-2 infected individuals (57). Interestingly, low-level replication has been suggested to occur in aviraemic HIV-2 infected individuals (52, 54), which could provide antigenic stimulation to immune cells. In line with this, aviraemic HIV-2 infected individuals display signs of immunopathology (24, 25, 58-63). The observation that HIV-2 more potently triggers IFN responses in dendritic cells, mediated through sensing by cyclic GMP-AMP synthase (cGAS) and TRIM5 (64, 65), could further explain how low-level tissue replication could still be sufficient to induce immune exhaustion in infected individuals. Further research is needed to elucidate the relationship between HIV replication in specific tissue compartments, chronic inflammation, and HIV disease progression.

In addition to the observed associations between HIV infection and GIT and spleen proteins, proteins previously found to be highly expressed in non-haematopoietic cells (NRP2, TNC, RAD23B, and LGALSL) could be linked to CD4% levels, indicating that HIV disease progression may be associated with a broad systemic impact on several cell types and organs (43, 44). Indeed, the chronic inflammation induced by HIV infection is known to impact several different cell types (46, 47). Within the cardiovascular system, HIV-1 infection has been reported to induce fibrotic remodelling, smooth muscle cell proliferation, and endothelial dysfunction, but less is known about the impact of HIV-2 infection (46). Interestingly, although we observed a negative association between the level of the smooth muscle cell enriched protein TNC and CD4% and CD4% midpoint, TNC has previously been able to bind directly to HIV-1 Env and inhibit infection (57). Further, in HIV-1 infected individuals, markers of monocyte activation, persistent inflammation, and hypercoagulation have been associated with increased risk of cardiovascular disease (66). Interestingly, we observed increased levels of endothelial and monocyte enhanced proteins among HIV-1, but not HIV-2, infected individuals.

We used DIA-mass spectrometry to perform an in-depth profiling of the plasma proteome in HIV-1 infected, HIV-2 infected, and HIV seronegative individuals. The major strength of DIA-mass-spectrometry is the ability to detect hundreds of proteins in a non-targeted approach (19). This makes mass-spectrometry highly suitable for the discovery of novel biomarkers for HIV infection, and to improve our understanding of HIV disease pathogenesis (19, 67). Mass-spectrometry has recently been used to study multiorgan-dysregulation in the murine sepsis models, allowing for linkage between blood plasma and organs within the same species (23, 68). To the best of our knowledge, data on large-scale proteomic profiling of human organs are not available, preventing similar studies from being performed based on human plasma. However, in two recent studies, the authors utilized the publicly available GTEx database to create tissue-enriched transcriptional signatures to study the impact of SARS-CoV-2 infection (37, 69). In addition, two studies from the HPA program characterized tissue and single cell RNA expression, with the support of immunohistochemistry, to create tissue and single cell expression maps (43, 44, 70). In the current study, we performed the first analysis of the contribution of HIV-1 and HIV-2 infection on tissue and single-cell type engagement by utilising the GTEx and HPA program databases. A limitation of our approach was that the previous definition of tissue and cell type enhanced genes were defined by RNA sequencing, and not protein expression. However, several of the identified tissues and cell types found to be engaged, has also previously been reported to be involved in HIV pathogenesis (47, 71, 72), supporting the relevance of this approach.

In conclusion, our broad-scale DIA-mass-spectrometry-based proteomic profiling of plasma from HIV uninfected and infected individuals showed that plasma protein levels associated with specific tissues and cell types could distinguish faster from slower HIV disease progression. The finding that both HIV-1 and HIV-2 infected individuals display signs of tissue and cell type engagement further support our previous findings that the majority of HIV-2 infected individuals will develop AIDS (7). Moreover, our findings open for further research on how specific target cells and non-haematopoietic bystander cells are linked to HIV disease progression – both in general and specifically related to progression of HIV-1 and HIV-2 infections.

## ACKNOWLEGMENTS

The authors thank the members of the Sweden Guinea-Bissau Cohort Research (SWEGUB CORE) group, including Babetida N’Buna, Antonio J. Biague, Ansu Biai, Cidia Camara, Joakim Esbjörnsson, Marianne Jansson, Emil Johansson, Sara Karlson, Jacob Lindman, Patrik Medstrand, Fredrik Månsson, Hans Norrgren, Angelica A. Palm, Gülsen Özkaya Sahin, Zacarias José da Silva, Zsòfia Ilona Szojka, and Sten Wilhelmson, and are indebted to the staff of the Police Clinics and the National Public Health Laboratory (LNSP) in Bissau, Guinea-Bissau. The authors also thank Sven Kjellström (Department of Clinical Sciences, BioMS, Lund University, Sweden) for his kind support. Support by NBIS (National Bioinformatics Infrastructure Sweden) is also gratefully acknowledged.

This work was supported by funding from the Swedish Research Council (grant number 2020-06262 to J.E; 2016-02285, 2019-01439 to M.J.) and by The Swedish Fund for Research without Animal Experiments to MJ (N2019-0009, F2021-0010).

## Author contributions

*Conceptualization:* Emil Johansson, Marianne Jansson, Joakim Esbjörnsson*: Data curation:* Emil Johansson*: Formal analysis*: Emil Johansson, Mun-Gwan Hong, Eva Freyhult. *Funding acquisition*: Marianne Jansson, Joakim Esbjörnsson*: Investigation*: Malin Neptin, Sara Karlsson, Melinda Rezeli, Marianne Jansson, Joakim Esbjörnsson*: Methodology:* Melinda Rezeli, Emil Johansson, Jamirah Nazziwa, Marianne Jansson, Joakim Esbjörnsson. *Project administration:* Emil Johansson, Marianne Jansson, Joakim Esbjörnsson*: Resources:* Jacob Lindman, Antonio Biague, Angelica Palm, Fredrik Månsson, Hans Norrgren, Marianne Jansson, Joakim Esbjörnsson*: Software:* Emil Johansson, Jamirah Nazziwa, Eva Freyhult, Mun-Gwan Hong*: Supervision*: Marianne Jansson, Joakim Esbjörnsson*: Validation:* Emil Johansson, Melinda Rezeli, Mun-Gwan Hong, Eva Freyhult: *Visualisation*: Emil Johansson. *Development of original draft*: Emil Johansson, Marianne Jansson, Joakim Esbjörnsson. *Review and editing*: All authors

## Conflicts of interest

The authors or their institutions declare no competing financial interests, and did not at any time receive payment or services from a third party (government, commercial, private foundation, etc.) for any aspect of the submitted work (including data monitoring board, study design, manuscript preparation, statistical analysis, etc.). The authors have no patents, whether planned, pending or issued, broadly relevant to the work.

## SUPPLEMENTARY MATERIAL

**Figure S1.**
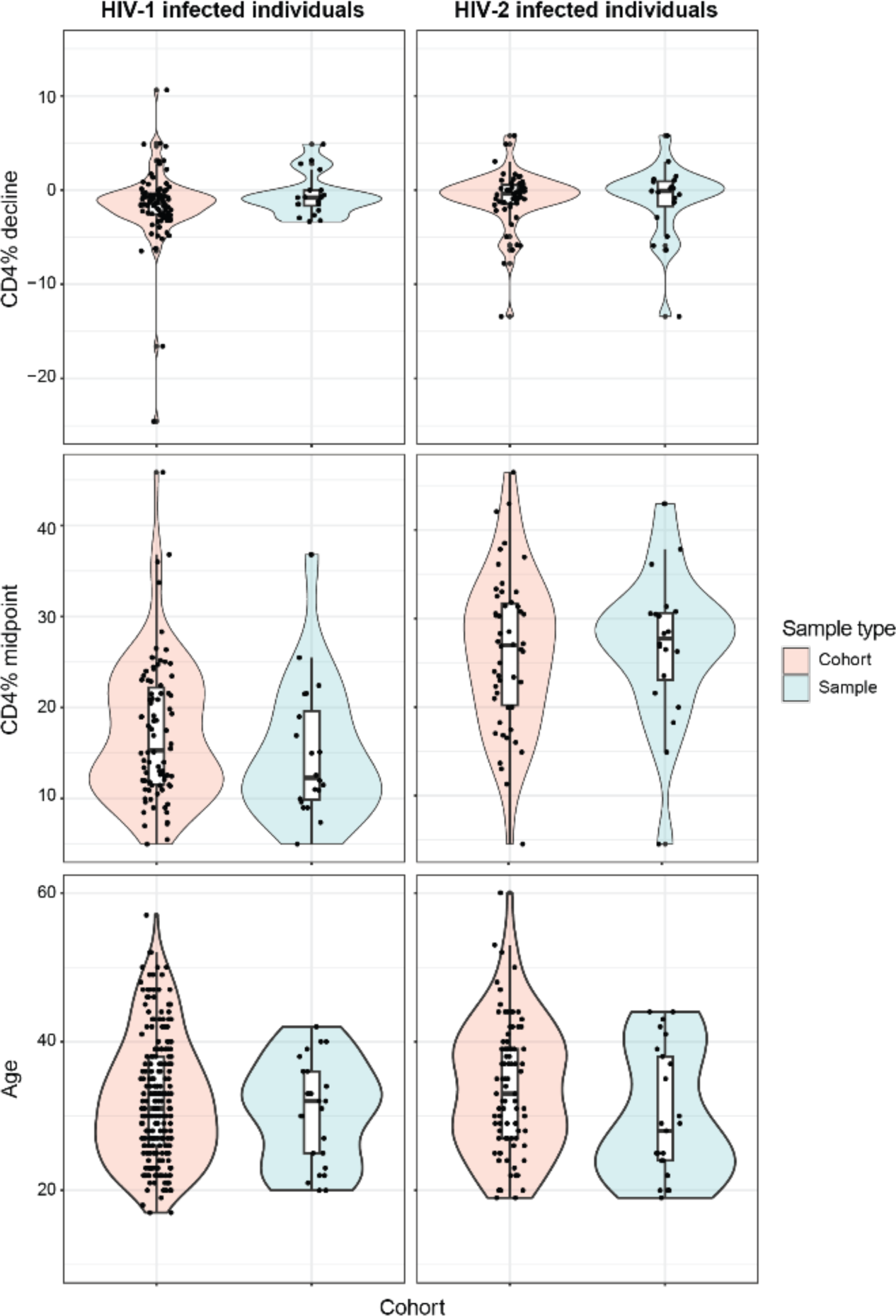
Included study participants are representative of the cohort. Violin plots visualising CD4% decline rate, CD4% midpoint, and age at first sample donation for included HIV infected individuals and all of the HIV-1 or HIV-2 seroconverting individuals in the occupational cohort. No significant difference was observed between any groups when compared using a Mann-Whitney U test.

**Figure S2.**
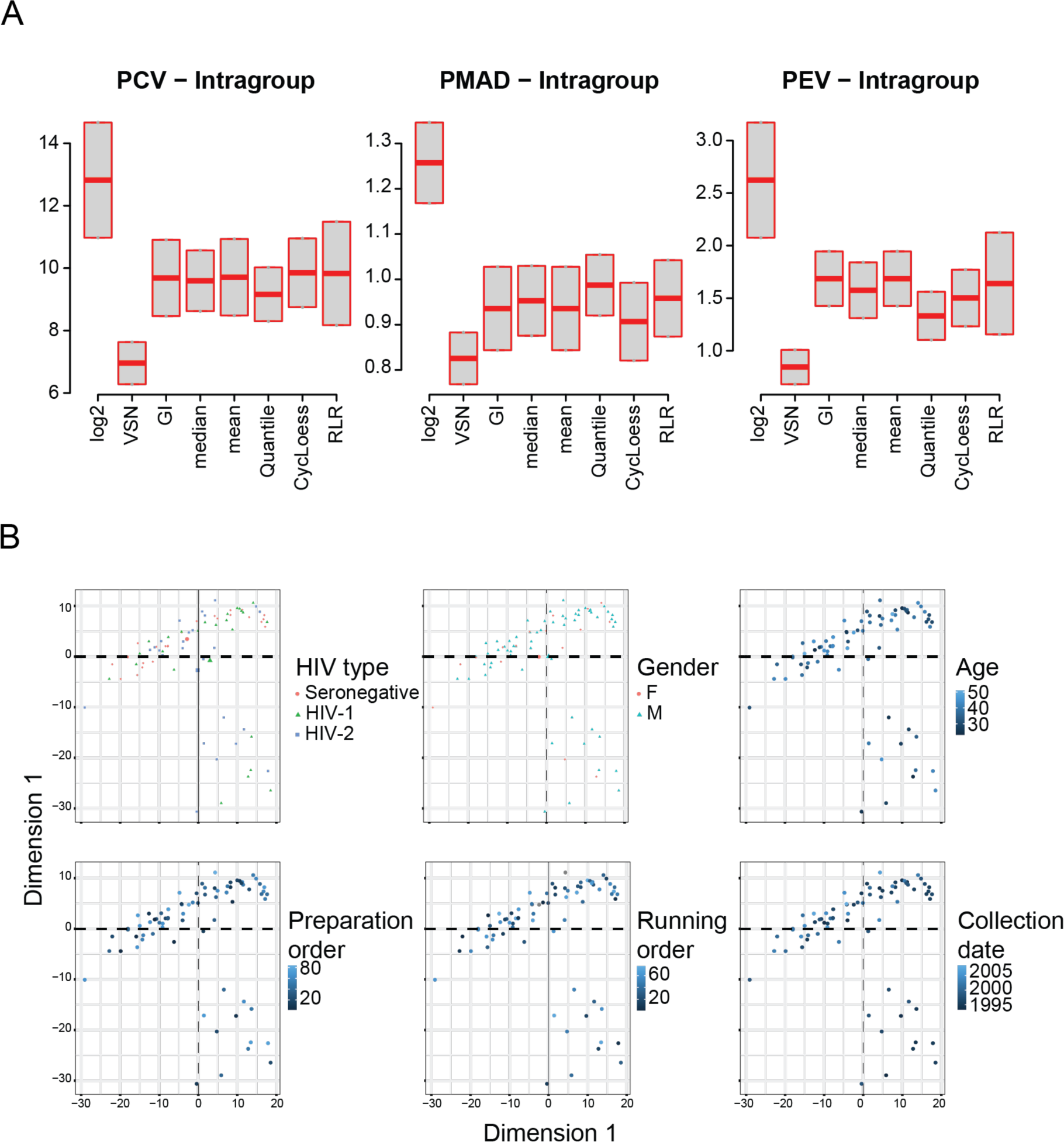
Quality control of proteomic data. A) Evaluation of Log2, variance stabilization normalization (VSN); global (GI), median, mean, quantile, cyclic loess (CycLoess), and robust linear regression (RLR) normalisation, using the NormalyzedDE package. HIV status groups were used as the group variable. B) The dimensionality of protein expression was reduced, by performing a principal component analysis (PCA). HIV status, sex, study participant age, sample preparation order, mass-spectrometry running order, and collection date was projected on the PCA plot. Abbreviations: PCV: mean of “intragroup CV of all replicate groups”, PMAD: mean of “intragroup median absolute deviation across all replicate groups”, PEV: mean of “intragroup pooled estimate of variance across all replicate groups”.

**Figure S3.**
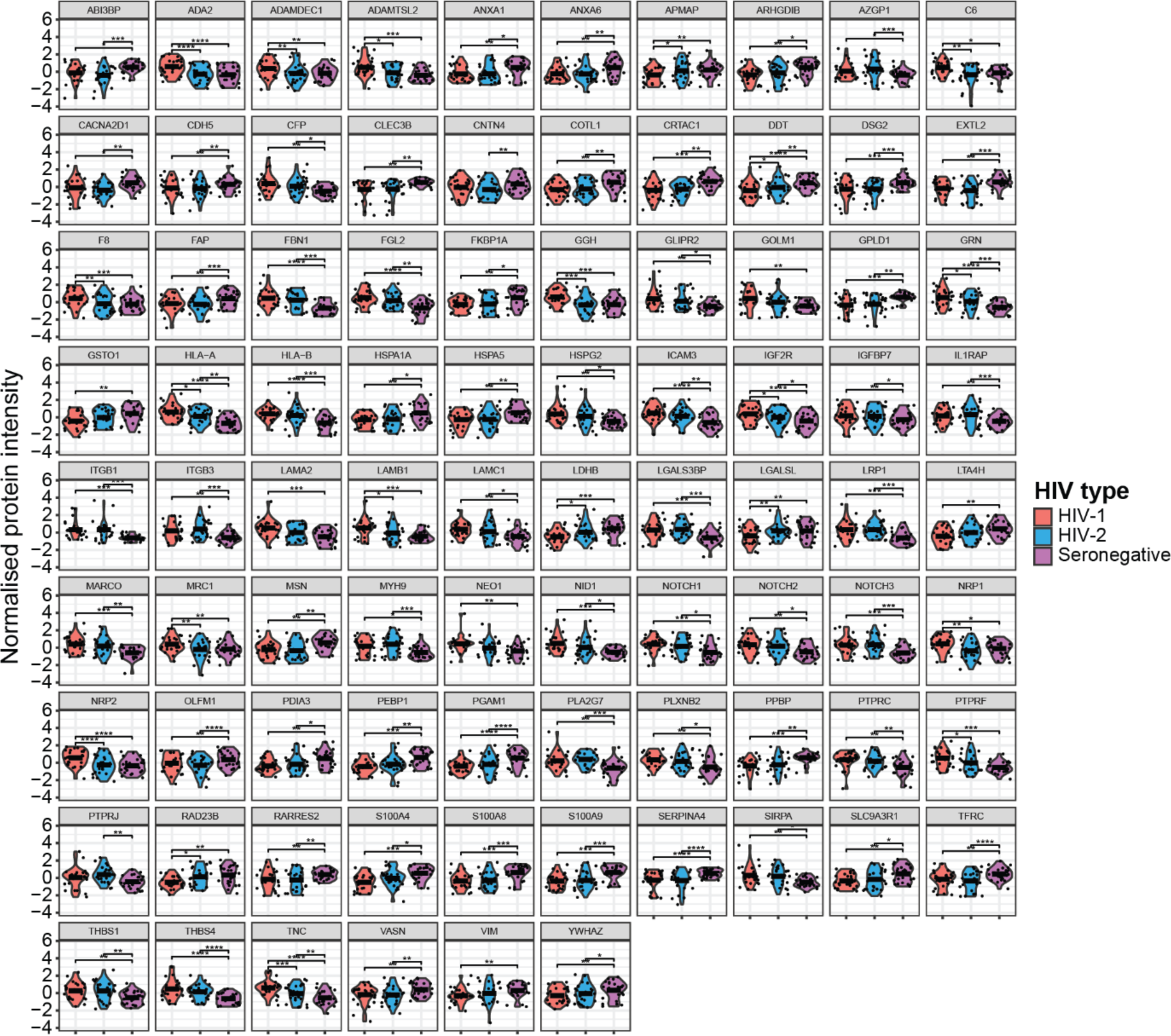
Plasma levels of 86 cell type enhanced proteins are differentially expressed between HIV status groups. A) ANOVA followed by Benjamini-Hochberg (BH) correction for multiple testing (p<0.05). Mean values are depicted in violin plots.

**Figure S4.**
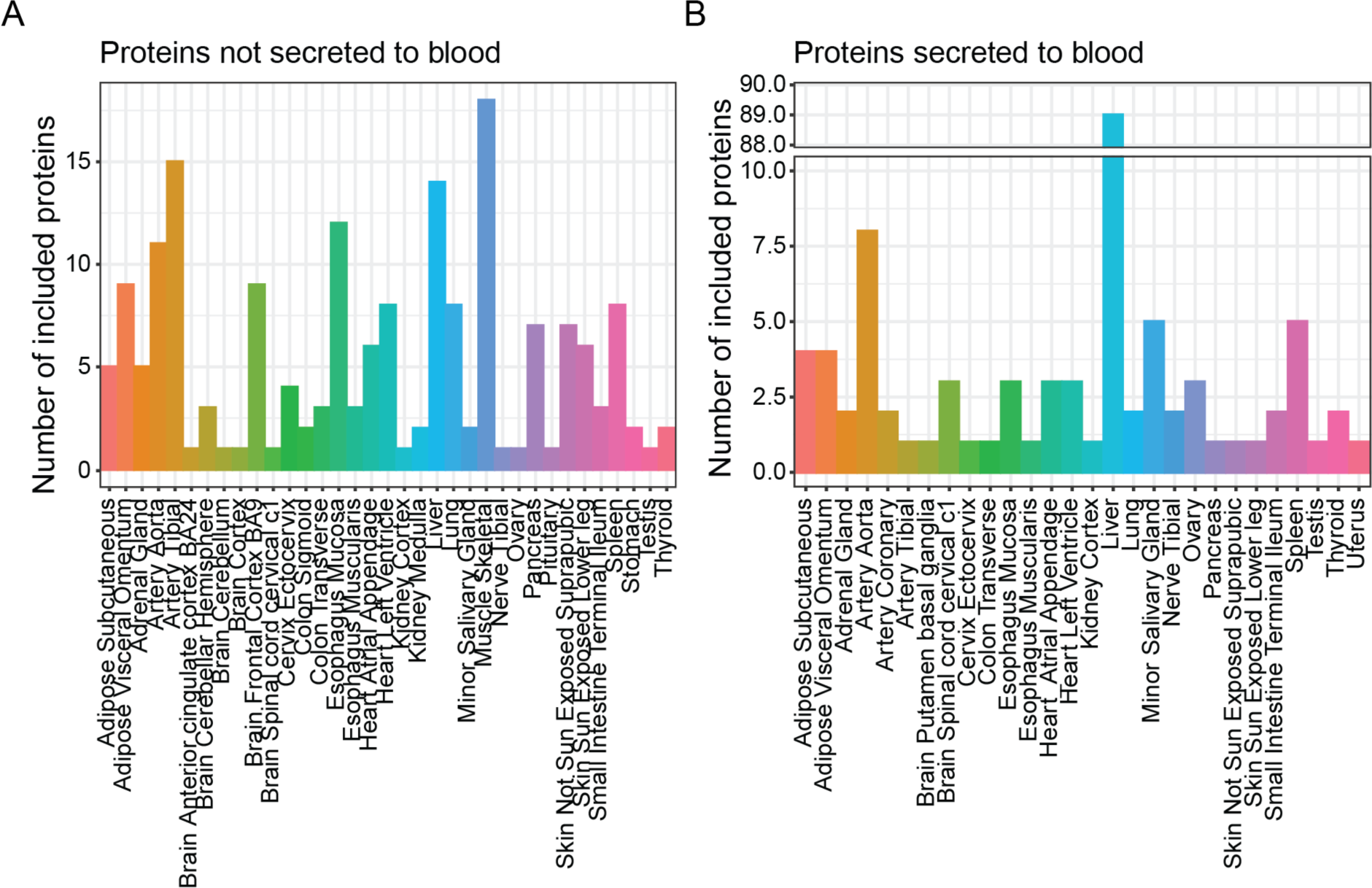
Variability in the number of proteins included in each tissue signature. Bar charts showing the number of A) leakage and B) secreted plasma proteins included in each tissue signature.

**Figure S5.**
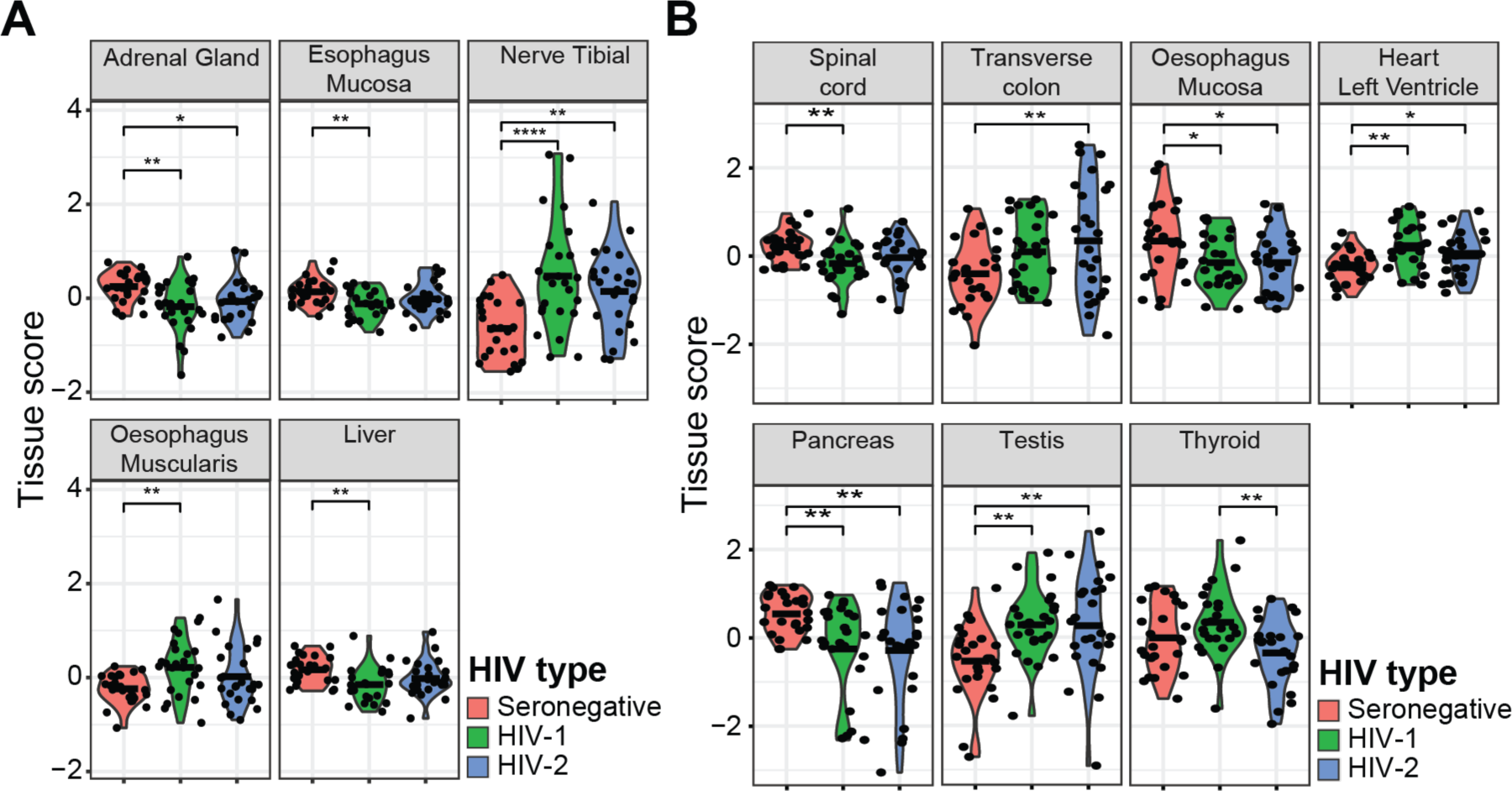
HIV-1 and HIV-2 infections induces alteration of tissue signatures obtained using leakage and secreted proteins. Violin plots of tissues signatures, consisting of A) leakage, or B) secreted plasma proteins. Groupwise comparisons were performed using an ANOVA test followed by (Benjamini-Hochberg (BH) correction for multiple testing, p<0.05). Mean values are depicted in violin plots.

**Figure S6.**
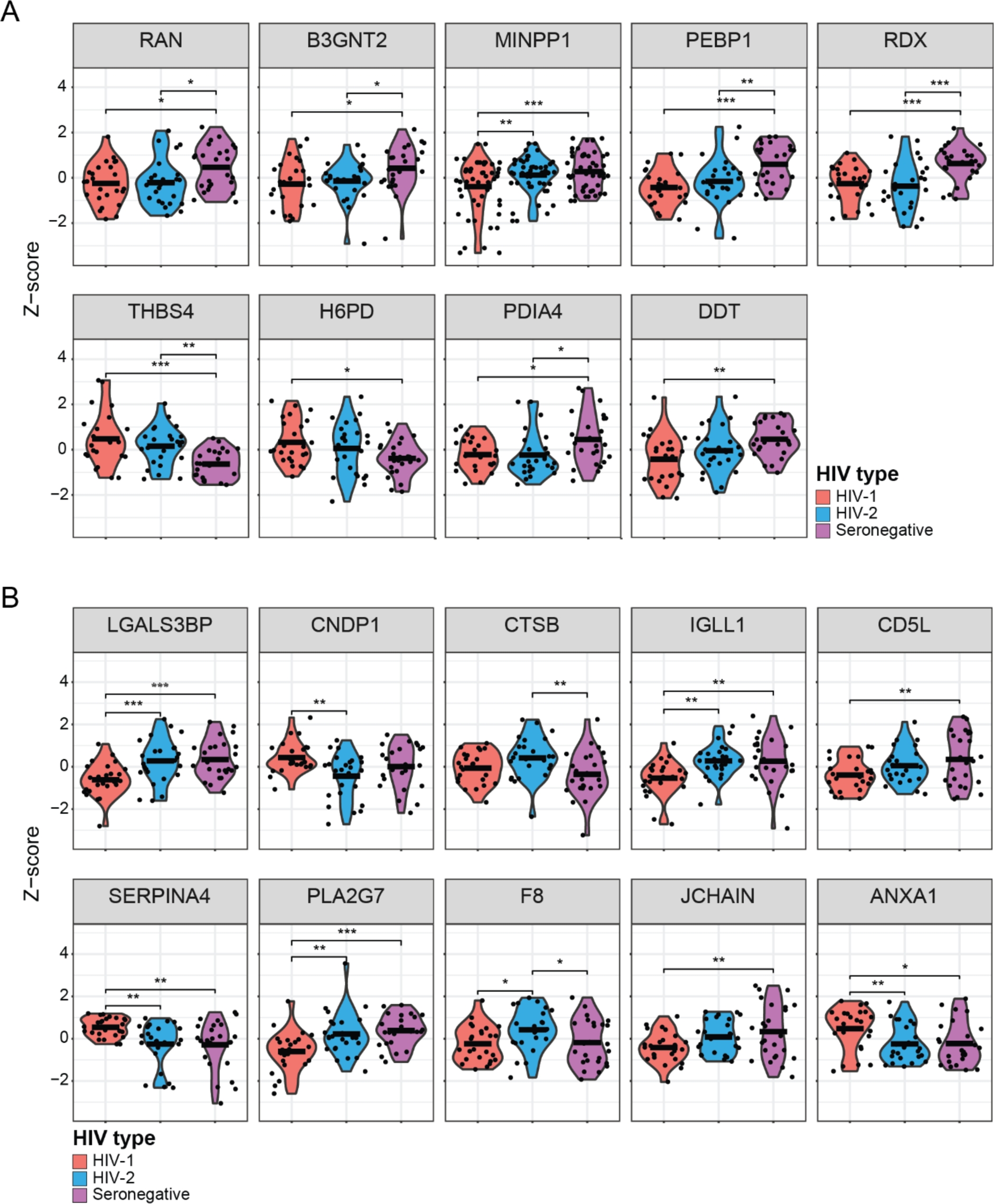
Both leakage and secreted plasma proteins are differentially expressed between HIV status groups. A) Leakage and B) secreted plasma proteins with z-score normalised protein intensity found to differ between HIV status groups. Groupwise comparisons were performed using an ANOVA test followed by Benjamini-Hochberg (BH) correction for multiple testing (p<0.05). Mean values are depicted in violin plots.

**Figure S7.**
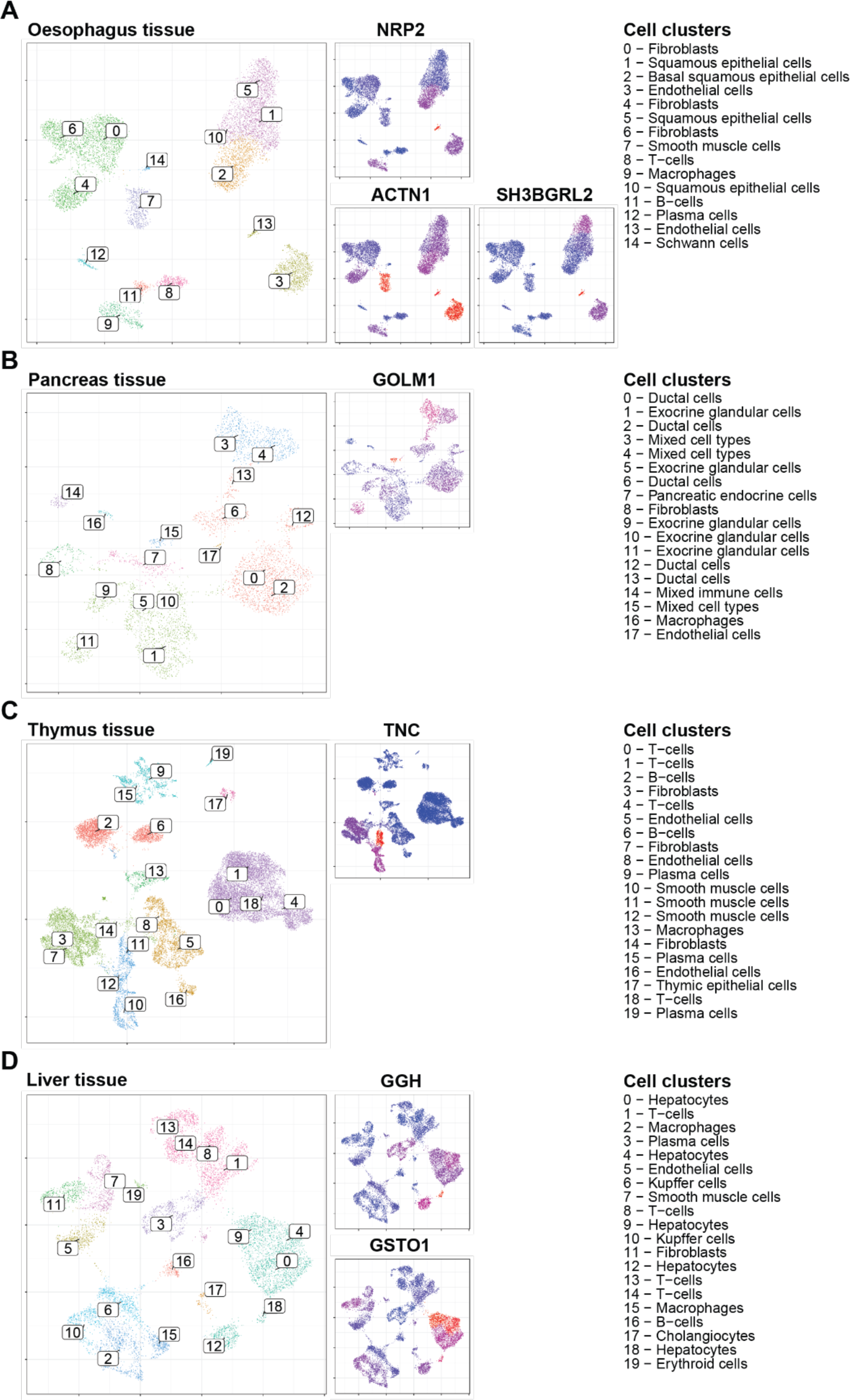
Tissue enriched proteins are expressed in a variety of cell types. Single cell RNA sequencing datasets from specified tissues were collected from the Human protein atlas program, and the previously determined cell clusters were projected on UMAP plots (large plots). In addition, the cluster-wise normalised transcript per million (nTPM) expression was projected on UMAP plots (small plots).

## Notes

### Competing Interest Statement

The authors have declared no competing interest.

### Author Declarations

The Ethical committee at Lund University gave ethical approval for this work The National Ethical Committee, Ministry of Public Health in Guinea-Bissau, gave ethical approval for this work

## REFERENCES

1. Berry N, Jaffar S, Schim van der Loeff M, Ariyoshi K, Harding E, N’Gom PT, et al. Low level viremia and high CD4% predict normal survival in a cohort of HIV type-2-infected villagers. AIDS Res Hum Retroviruses. 2002;18(16):1167–73.

2. van der Loeff MF, Larke N, Kaye S, Berry N, Ariyoshi K, Alabi A, et al. Undetectable plasma viral load predicts normal survival in HIV-2-infected people in a West African village. Retrovirology. 2010;7:46.

3. de Silva TI, Cotten M, Rowland-Jones SL. HIV-2: the forgotten AIDS virus. Trends Microbiol. 2008;16(12):588–95.

4. Poulsen AG, Aaby P, Larsen O, Jensen H, Naucler A, Lisse IM, et al. 9-year HIV-2-associated mortality in an urban community in Bissau, west Africa. Lancet. 1997;349(9056):911-4.

5. Rowland-Jones SL, Whittle HC. Out of Africa: what can we learn from HIV-2 about protective immunity to HIV-1? Nat Immunol. 2007;8(4):329–31.

6. Tchounga B, Ekouevi DK, Balestre E, Dabis F. Mortality and survival patterns of people living with HIV-2. Curr Opin HIV AIDS. 2016;11(5):537–44.

7. Esbjornsson J, Mansson F, Kvist A, da Silva ZJ, Andersson S, Fenyo EM, et al. Long-term follow-up of HIV-2-related AIDS and mortality in Guinea-Bissau: a prospective open cohort study. Lancet HIV. 2018;6(1):e25-e31.

8. Esbjornsson J, Mansson F, Lindman J, Rowland-Jones SL, Jansson M, Medstrand P, et al. New insights are game-changers in HIV-2 disease management - Authors’ reply. Lancet HIV. 2019;6(4):e214–e5.

9. de Silva TI, Aasa-Chapman M, Cotten M, Hue S, Robinson J, Bibollet-Ruche F, et al. Potent autologous and heterologous neutralizing antibody responses occur in HIV-2 infection across a broad range of infection outcomes. J Virol. 2012;86(2):930–46.

10. Duvall MG, Jaye A, Dong T, Brenchley JM, Alabi AS, Jeffries DJ, et al. Maintenance of HIV-specific CD4+ T cell help distinguishes HIV-2 from HIV-1 infection. J Immunol. 2006;176(11):6973–81.

11. Duvall MG, Precopio ML, Ambrozak DA, Jaye A, McMichael AJ, Whittle HC, et al. Polyfunctional T cell responses are a hallmark of HIV-2 infection. Eur J Immunol. 2008;38(2):350–63.

12. Karlsson I, Tingstedt JL, Sahin GO, Hansen M, Szojka Z, Buggert M, et al. Cross-Reactive Antibodies With the Capacity to Mediate HIV-1 Envelope Glycoprotein-Targeted Antibody-Dependent Cellular Cytotoxicity Identified in HIV-2-Infected Individuals. J Infect Dis. 2019;219(11):1749–54.

13. Kong R, Li H, Bibollet-Ruche F, Decker JM, Zheng NN, Gottlieb GS, et al. Broad and potent neutralizing antibody responses elicited in natural HIV-2 infection. J Virol. 2012;86(2):947–60.

14. Leligdowicz A, Onyango C, Yindom LM, Peng Y, Cotten M, Jaye A, et al. Highly avid, oligoclonal, early-differentiated antigen-specific CD8+ T cells in chronic HIV-2 infection. Eur J Immunol. 2010;40(7):1963–72.

15. Ozkaya Sahin G, Holmgren B, da Silva Z, Nielsen J, Nowroozalizadeh S, Esbjornsson J, et al. Potent intratype neutralizing activity distinguishes human immunodeficiency virus type 2 (HIV-2) from HIV-1. J Virol. 2012;86(2):961-71.

16. Ozkaya Sahin G, Holmgren B, Sheik-Khalil E, da Silva Z, Nielsen J, Nowroozalizadeh S, et al. Effect of complement on HIV-2 plasma antiviral activity is intratype specific and potent. J Virol. 2013;87(1):273-81.

17. Berry N, Ariyoshi K, Jaffar S, Sabally S, Corrah T, Tedder R, et al. Low peripheral blood viral HIV-2 RNA in individuals with high CD4 percentage differentiates HIV-2 from HIV-1 infection. J Hum Virol. 1998;1(7):457–68.

18. Esbjornsson J, Jansson M, Jespersen S, Mansson F, Honge BL, Lindman J, et al. HIV-2 as a model to identify a functional HIV cure. AIDS Res Ther. 2019;16(1):24.

19. Donnelly MR, Ciborowski P. Proteomics, biomarkers, and HIV-1: A current perspective. Proteomics Clin Appl. 2016;10(2):110–25.

20. Thul PJ, Akesson L, Wiking M, Mahdessian D, Geladaki A, Ait Blal H, et al. A subcellular map of the human proteome. Science. 2017;356(6340).

21. Uhlen M, Fagerberg L, Hallstrom BM, Lindskog C, Oksvold P, Mardinoglu A, et al. Proteomics. Tissue-based map of the human proteome. Science. 2015;347(6220):1260419.

22. Happonen L, Hauri S, Svensson Birkedal G, Karlsson C, de Neergaard T, Khakzad H, et al. A quantitative Streptococcus pyogenes-human protein-protein interaction map reveals localization of opsonizing antibodies. Nat Commun. 2019;10(1):2727.

23. Malmstrom E, Kilsgard O, Hauri S, Smeds E, Herwald H, Malmstrom L, et al. Large-scale inference of protein tissue origin in gram-positive sepsis plasma using quantitative targeted proteomics. Nat Commun. 2016;7:10261.

24. Buggert M, Frederiksen J, Lund O, Betts MR, Biague A, Nielsen M, et al. CD4+ T cells with an activated and exhausted phenotype distinguish immunodeficiency during aviremic HIV-2 infection. AIDS. 2016;30(16):2415–26.

25. Scharf L, Pedersen CB, Johansson E, Lindman J, Olsen LR, Buggert M, et al. Inverted CD8 T-Cell Exhaustion and Co-Stimulation Marker Balance Differentiate Aviremic HIV-2-Infected From Seronegative Individuals. Front Immunol. 2021;12:744530.

26. Zhang H, Orti G, Du Q, He J, Kankasa C, Bhat G, et al. Phylogenetic and phenotypic analysis of HIV type 1 env gp120 in cases of subtype C mother-to-child transmission. AIDS Res Hum Retroviruses. 2002;18(18):1415–23.

27. Pham TV, Henneman AA, Jimenez CR. iq: an R package to estimate relative protein abundances from ion quantification in DIA-MS-based proteomics. Bioinformatics. 2020;36(8):2611–3.

28. Smilde AK, van der Werf MJ, Bijlsma S, van der Werff-van der Vat BJC, Jellema RH. Fusion of Mass Spectrometry-Based Metabolomics Data. Analytical Chemistry. 2005;77(20):6729–36.

29. Palm AA, Lemey P, Jansson M, Mansson F, Kvist A, Szojka Z, et al. Low Postseroconversion CD4(+) T-cell Level Is Associated with Faster Disease Progression and Higher Viral Evolutionary Rate in HIV-2 Infection. mBio. 2019;10(1).

30. Boswell MT, Nazziwa J, Kuroki K, Palm A, Karlson S, Mansson F, et al. Intrahost evolution of the HIV-2 capsid correlates with progression to AIDS. Virus Evol. 2022;8(2):veac075.

31. Willforss J, Chawade A, Levander F. NormalyzerDE: Online Tool for Improved Normalization of Omics Expression Data and High-Sensitivity Differential Expression Analysis. J Proteome Res. 2019;18(2):732–40.

32. Lê S, Josse J, Husson F. FactoMineR: An R Package for Multivariate Analysis. Journal of Statistical Software. 2008;25(1):1–18.

33. R Core Team (2022). R: A language and environment for statistical computing. R Foundation for Statistical Computing, Vienna, Austria. [Available from: https://www.R-project.org/ [accessed on 03-04-22].

34. Ritchie ME, Phipson B, Wu D, Hu Y, Law CW, Shi W, et al. limma powers differential expression analyses for RNA-sequencing and microarray studies. Nucleic Acids Research. 2015;43(7):e47-e.

35. Wu T, Hu E, Xu S, Chen M, Guo P, Dai Z, et al. clusterProfiler 4.0: A universal enrichment tool for interpreting omics data. Innovation (Camb). 2021;2(3):100141.

36. Yu G, Wang LG, Han Y, He QY. clusterProfiler: an R package for comparing biological themes among gene clusters. Omics. 2012;16(5):284–7.

37. Arthur L, Esaulova E, Mogilenko DA, Tsurinov P, Burdess S, Laha A, et al. Cellular and plasma proteomic determinants of COVID-19 and non-COVID-19 pulmonary diseases relative to healthy aging. Nature Aging. 2021;1(6):535–49.

38. Kassambara A KM, Biecek P. survminer: Drawing Survival Curves using ‘ggplot2’ 2021 [Available from: https://CRAN.R-project.org/package=survminer [Accessed 2023-02-16].

39. T T. A Package for Survival Analysis in R 2023 [Available from: https://CRAN.R-project.org/package=survival [Accessed 2023-02-16].

40. Wickham H. ggplot2: Elegant Graphics for Data Analysis. New York, NY: Springer; 2016.

41. Blighe K, Rana S, M L. EnhancedVolcano: Publication-ready volcano plots with enhanced colouring and labeling. 2018.

42. The proteins actively secreted to human blood - Human Protein Atlas [Available from: https://www.proteinatlas.org/humanproteome/blood+protein/secreted+to+blood [Accessed February 8, 2023].

43. Atlas HP. [Available from: proteinatlas.org [Accessed 2023-02-08].

44. Karlsson M, Zhang C, Méar L, Zhong W, Digre A, Katona B, et al. A single-cell type transcriptomics map of human tissues. Science Advances. 2021;7(31):eabh2169.

45. Valentin A, Albert J, Fenyö EM, Asjö B. Dual tropism for macrophages and lymphocytes is a common feature of primary human immunodeficiency virus type 1 and 2 isolates. J Virol. 1994;68(10):6684–9.

46. Teer E, Dominick L, Mukonowenzou NC, Essop MF. HIV-Related Myocardial Fibrosis: Inflammatory Hypothesis and Crucial Role of Immune Cells Dysregulation. Cells. 2022;11(18):2825.

47. Mazzuti L, Turriziani O, Mezzaroma I. The Many Faces of Immune Activation in HIV-1 Infection: A Multifactorial Interconnection. Biomedicines. 2023;11(1).

48. Brenchley JM, Price DA, Schacker TW, Asher TE, Silvestri G, Rao S, et al. Microbial translocation is a cause of systemic immune activation in chronic HIV infection. Nat Med. 2006;12(12):1365–71.

49. Epple HJ, Allers K, Tröger H, Kühl A, Erben U, Fromm M, et al. Acute HIV infection induces mucosal infiltration with CD4+ and CD8+ T cells, epithelial apoptosis, and a mucosal barrier defect. Gastroenterology. 2010;139(4):1289–300.

50. Cassol E, Malfeld S, Mahasha P, van der Merwe S, Cassol S, Seebregts C, et al. Persistent microbial translocation and immune activation in HIV-1-infected South Africans receiving combination antiretroviral therapy. J Infect Dis. 2010;202(5):723–33.

51. Nowroozalizadeh S, Månsson F, da Silva Z, Repits J, Dabo B, Pereira C, et al. Microbial translocation correlates with the severity of both HIV-1 and HIV-2 infections. J Infect Dis. 2010;201(8):1150-4.

52. Fernandes SM, Pires AR, Matoso P, Ferreira C, Nunes-Cabaco H, Correia L, et al. HIV-2 infection is associated with preserved GALT homeostasis and epithelial integrity despite ongoing mucosal viral replication. Mucosal Immunol. 2018;11(1):236–48.

53. Wong JK, Yukl SA. Tissue reservoirs of HIV. Curr Opin HIV AIDS. 2016;11(4):362–70.

54. Soares RS, Tendeiro R, Foxall RB, Baptista AP, Cavaleiro R, Gomes P, et al. Cell-associated viral burden provides evidence of ongoing viral replication in aviremic HIV-2-infected patients. J Virol. 2011;85(5):2429–38.

55. Lucas SB, Hounnou A, Peacock C, Beaumel A, Djomand G, N’Gbichi J-M, et al. The mortality and pathology of HIV infection in a West African city. AIDS. 1993;7(12):1569–79.

56. Martinez-Steele E, Awasana AA, Corrah T, Sabally S, van der Sande M, Jaye A, et al. Is HIV-2-induced AIDS different from HIV-1-associated AIDS? Data from a West African clinic. AIDS. 2007;21(3):317–24.

57. Nyamweya S, Hegedus A, Jaye A, Rowland-Jones S, Flanagan KL, Macallan DC. Comparing HIV-1 and HIV-2 infection: Lessons for viral immunopathogenesis. Rev Med Virol. 2013;23(4):221–40.

58. Bachle SM, Malone DF, Buggert M, Karlsson AC, Isberg PE, Biague AJ, et al. Elevated levels of invariant natural killer T-cell and natural killer cell activation correlate with disease progression in HIV-1 and HIV-2 infections. AIDS. 2016;30(11):1713–22.

59. Cavaleiro R, Baptista AP, Soares RS, Tendeiro R, Foxall RB, Gomes P, et al. Major depletion of plasmacytoid dendritic cells in HIV-2 infection, an attenuated form of HIV disease. PLoS Pathog. 2009;5(11):e1000667.

60. Cavaleiro R, Tendeiro R, Foxall RB, Soares RS, Baptista AP, Gomes P, et al. Monocyte and myeloid dendritic cell activation occurs throughout HIV type 2 infection, an attenuated form of HIV disease. J Infect Dis. 2013;207(11):1730–42.

61. Palm AA, Veerla S, Lindman J, Isberg P-E, Johansson E, Biague A, et al. Interferon Alpha-Inducible Protein 27 Expression Is Linked to Disease Severity in Chronic Infection of Both HIV-1 and HIV-2. Frontiers in Virology. 2022;2.

62. Tendeiro R, Fernandes S, Foxall RB, Marcelino JM, Taveira N, Soares RS, et al. Memory B-cell depletion is a feature of HIV-2 infection even in the absence of detectable viremia. AIDS. 2012;26(13):1607–17.

63. Sousa AE, Carneiro J, Meier-Schellersheim M, Grossman Z, Victorino RM. CD4 T cell depletion is linked directly to immune activation in the pathogenesis of HIV-1 and HIV-2 but only indirectly to the viral load. J Immunol. 2002;169(6):3400–6.

64. Lahaye X, Satoh T, Gentili M, Cerboni S, Conrad C, Hurbain I, et al. The Capsids of HIV-1 and HIV-2 Determine Immune Detection of the Viral cDNA by the Innate Sensor cGAS in Dendritic Cells. Immunity. 2013;39(6):1132–42.

65. Zuliani-Alvarez L, Govasli ML, Rasaiyaah J, Monit C, Perry SO, Sumner RP, et al. Evasion of cGAS and TRIM5 defines pandemic HIV. Nat Microbiol. 2022;7(11):1762–76.

66. So-Armah K, Benjamin LA, Bloomfield GS, Feinstein MJ, Hsue P, Njuguna B, et al. HIV and cardiovascular disease. Lancet HIV. 2020;7(4):e279–e93.

67. Macklin A, Khan S, Kislinger T. Recent advances in mass spectrometry based clinical proteomics: applications to cancer research. Clinical Proteomics. 2020;17(1):17.

68. Toledo AG, Golden G, Campos AR, Cuello H, Sorrentino J, Lewis N, et al. Proteomic atlas of organ vasculopathies triggered by Staphylococcus aureus sepsis. Nature Communications. 2019;10(1):4656.

69. Filbin MR, Mehta A, Schneider AM, Kays KR, Guess JR, Gentili M, et al. Longitudinal proteomic analysis of severe COVID-19 reveals survival-associated signatures, tissue-specific cell death, and cell-cell interactions. Cell Rep Med. 2021;2(5):100287.

70. Uhlén M, Fagerberg L, Hallström BM, Lindskog C, Oksvold P, Mardinoglu A, et al. Tissue-based map of the human proteome. Science. 2015;347(6220):1260419.

71. Perkins MV, Joseph SB, Dittmer DP, Mackman N. Cardiovascular Disease and Thrombosis in HIV Infection. Arterioscler Thromb Vasc Biol. 2023;43(2):175–91.

72. Joseph J, Daley W, Lawrence D, Lorenzo E, Perrin P, Rao VR, et al. Role of macrophages in HIV pathogenesis and cure: NIH perspectives. J Leukoc Biol. 2022;112(5):1233–43.

